# Health and economic effects of introducing single-dose human papillomavirus vaccination in India

**DOI:** 10.1101/2023.04.14.23288563

**Authors:** TM de Carvalho, I Man, D Georges, LR Saraswati, P Bhandari, I Kataria, M Siddiqui, R Muwonge, E Lucas, R Sankaranarayanan, P Basu, J Berkhof, JA Bogaards, I Baussano

**Author notes:** Author to whom correspondence should be addressed Correspondence to: Dr Tiago M de Carvalho, Department of Epidemiology and Data Science, Amsterdam UMC, The Netherlands. Authors share co-senior authorship (JA Bogaards, I Baussano).

## Abstract

**Background:** Cervical cancer is a major public health problem in India, where access to prevention programmes is low. The World Health Organization-Strategic Advisory Group of Experts recently updated their recommendation for human papillomavirus (HPV) vaccination to include a single-dose option in addition to the two-dose option, which could make HPV vaccination programmes easier to implement and more affordable.

**Methods:** We combined projections from a type-specific HPV transmission model and a cancer progression model to assess the health and economic effects of HPV vaccination at national and state-level in India. The models used national and state-specific Indian demographic, epidemiological and cost data, and single-dose vaccine efficacy and immunogenicity data from the IARC India vaccine trial with 10-year follow-up. We compared single- and two-dose HPV vaccination for a range of plausible scenarios regarding single-dose vaccine protection, coverage and catch-up.

**Results:** Under the base-case scenario of life-long protection of single-dose vaccination in 10-year-old girls with 90% coverage, the incremental cost-effectiveness ratio (ICER) of nationwide vaccination relative to no vaccination was $405 per DALY averted and lay below an opportunity-cost based threshold of 30% Indian GDP per capita in each state (state-specific ICER range: $67 to $593 per DALY averted). The ICER of two-dose vaccination versus no vaccination and versus single-dose vaccination was $1403 and minimum $2279 per DALY averted, respectively.

**Conclusions:** Nationwide introduction of single-dose HPV vaccination in India is highly likely to be cost-effective whereas extending the number of doses from one to two would have a less favourable profile.

**Funding:** Bill & Melinda Gates Foundation.

**What is already known in this topic:** In 2020, the World Health Organization (WHO) launched a global call for elimination of cervical cancer as a public health problem, of which HPV vaccination is a key pillar. However, access to HPV vaccination in India is still very low.

In April 2022, the WHO Strategic Advisory Group of Experts (SAGE) issued a recommendation for countries to update their dosing schedules to include a single-dose option. Single-dose HPV vaccination is likely to be more affordable and would greatly facilitate the implementation of HPV vaccination.

The key questions for India are whether, with a realistic cost-effectiveness threshold (30% GDP per capita), single-dose HPV vaccination would be a cost-effective intervention; and whether two-dose vaccination could still be affordable and worthwhile compared to a single-dose schedule, given the uncertainty in its initial efficacy and long-term protection.

**What this study adds:** We used state-specific cancer incidence and locally collected cost data and built plausible vaccination efficacy scenarios based on the IARC India trial to inform the cost-effectiveness estimates.

Single-dose vaccination in India would be cost-effective under a cost-effectiveness threshold of 30% of the Indian GDP per capita and the annual budget impact would be less than 10% of the cost of the current Indian universal childhood vaccination programme.

Even though there was substantial heterogeneity, we confirmed that single-dose vaccination would be cost-effective across all Indian states.

Catch-up single-dose vaccination to age 15 or 20 is a cost-effective strategy. However, the decision to implement catch-up will depend on the willingness of the health authorities to support a higher initial investment. We found two-dose vaccination to have a less favourable cost-effectiveness profile.

**How this study might affect research practice and policy:** Single-dose vaccination achieved a better balance between health benefits and financial burden than two-dose vaccination, even after taking into account uncertainty in the level of protection provided by single-dose HPV vaccination.

Our results could be used by Indian health authorities at the national and state-level to inform their decision and planning of the implementation of HPV vaccination in India and could convey several lessons for other low and middle income countries.

## INTRODUCTION

Cervical cancer is a major public health problem. Globally, an estimated 604,000 women were diagnosed with cervical cancer, and 342,000 died from cervical cancer in 2020.^1^ Most of the cancer burden is located in low- and middle-income countries (LMICs), and in particular, one sixth of the future cervical cancer cases will be diagnosed in India.^2^ HPV vaccination has a high efficacy for preventing cervical cancer,^3,4^ and is a key component of the World Health Organization (WHO) strategy for worldwide elimination of cervical cancer as a public health problem.^5^ The prospect of introducing HPV vaccination into the Indian national immunisation programme (NIP) has improved following the recent marketing authorisation granted to a nationally produced vaccine.^6^ However, at its current cost, implementing two-dose HPV vaccination still requires a large investment, while the health benefits could take decades to accrue.^7^

In April 2022, the WHO Strategic Advisory Group of Experts (SAGE) recommended countries to update their dosing schedules to include a single-dose option.^8^ This recommendation was based on data from the IARC India vaccine trial,^9^ the randomised controlled trial in Kenya,^10^ and other evidence demonstrating high efficacy of single-dose protection against persistent HPV 16/18 infection, comparable to two doses. Single-dose vaccination could make HPV vaccination more affordable. However, there is uncertainty regarding the protection afforded by single-dose vaccination beyond 10 years,^11^ since antibody levels induced by single-dose vaccination appear to be lower than by multiple-dose vaccination.

In this study, we derived a comprehensive projection of the health and economic effects of vaccinating girls against HPV in India. As decisions on introducing HPV vaccination and on the required dose schedule must be made well ahead of obtaining information on long-term single-dose protection, this modelling analysis aimed to help inform Indian policymakers in their decisions-making on cervical cancer prevention by assessing the cost-effectiveness of single-dose and two-dose HPV vaccination, under a more stringent cost-effectiveness threshold suggested for LMICSs (30% GDP per capita) in recent health economics literature. ^12-14^ For this, we used 10-year follow-up data from the IARC India vaccine trial ^9^ to derive single-dose vaccine efficacy and duration of protection scenarios, and demographic, epidemiological and cost data on a state-specific level, to obtain nationwide and state-specific projections for India.

## METHODS

### Patient Involvement

No patients were involved in the design and conduct of this research.

### Modelling Approach

We combined an HPV transmission model and a cervical cancer progression model to estimate the health and economic effects of HPV vaccination. The first model, EpiMetHeos, extends an open source framework (EpiModel) for the simulation of dynamic contact networks,^15^ and describes transmission of 13 type-specific HPV infection types in India. This model is extensively described in Man et al.^16^ Using HPV incidence estimated from EpiMetHeos, the second model simulates the progression from HPV infection to cervical intraepithelial neoplasia (CIN), cervical cancer and death, and is described in the supplementary appendix.

We assumed that while HPV infection incidence can be country- or state-specific, the natural history of HPV infection to cervical cancer does not vary across countries. This is a biologically plausible assumption for women who are not immunocompromised and is commonly adopted in multi-country modelling studies.^17,18^ Natural history parameters were estimated in previous publications based on longitudinal follow-up in the POBASCAM study,^19,20^ and by statistically linking the age distributions of CIN2/3 and cervical cancer, based on data from the Dutch National Pathology Databank (PALGA) and the Dutch Cancer Registry (IKNL).^21^ Further details are provided in Online Supplement Table A.1 and Online Supplement Figure A.1.

### Model adaptation to India

We calibrated the model to Indian national and subnational data to derive nationwide and state-specific impact projections of HPV vaccination. Since not all Indian states had high-quality data on cervical cancer incidence, we derived state-level projections in three steps: 1) we identified clusters of states with high or low cancer incidence; 2) we calibrated EpiMetHeos to type-specific HPV prevalence and sexual behaviour data from a representative high (Tamil Nadu) and low (West Bengal) cancer incidence state (Online Supplement Figure A.2); 3) we used the resulting projections to extrapolate cancer incidence to other states within the same cluster. Details about the calibration and clustering are provided elsewhere.^22,23^

Furthermore, we adjusted the cancer detection rates to match the observed cervical cancer stage distribution in India, and we used key Indian demographic and epidemiological data, including background mortality rate, age distribution and hysterectomy rate. Finally, we extracted 5-year Indian cancer survival based on a literature review (Online Supplement Tables A.2-A.4). Throughout this study, we assumed that no screening took place in India, which is justified by the current very low uptake of opportunistic screening.^24^

### Cost Data Collection

We collected Indian data on cervical cancer treatment costs from four hospitals and from the literature (Online Supplement Tables A.5-A.8).^25^ Cost of the HPV vaccine was set at the GAVI price ($USD 4.5 per dose). Costs of vaccination programme implementation and delivery were collected for the state of Sikkim, where a pilot HPV programme was implemented. In order to extrapolate these costs to the rest of India, we used state-level delivery cost estimates of the childhood vaccination programme,^26^ and assumed that the relative differences in delivery costs between states were the same for these programmes (Online Supplement Tables A.9-A.12).

### Vaccination Scenarios

In the base-case scenario, we set vaccination age at 10 years and coverage at 90% as per WHO recommendations. We evaluated both a single-dose and a two-dose schedule. For the two-dose schedule we assumed no drop-out between the first and second dose. We derived four assumptions of vaccine efficacy, based on the lower bound of the vaccine efficacy estimate of the IARC India vaccine trial ^9^, and duration of single-dose protection, informed by trial immunogenicity data, based on the time until antibody levels of HPV 16/18 have decreased below different detection thresholds. We refer to Man et al.^16^ for details on the construction of these assumptions. In assumption A (used in the base-case scenario) vaccine efficacies under single-dose schedule were assumed to be 95% for HPV 16/18, 9% for HPV 31/33/45, and 0% for the remaining oncogenic HPV types, with lifetime duration of vaccine protection. Assumptions B, C, and D corresponded to a remaining efficacy 20 years after vaccination of approximately 80%, 75% and 65% for HPV 16/18, respectively. Additionally, assumptions C and D had a lower initial efficacy equal to 90% and 85% for HPV16 and 85% and 55% for HPV18, respectively (Online Supplement Table A.13).

In combination with vaccine efficacy and waning assumptions A–D, we evaluated multiple levels of vaccination coverage (60%–100%). Finally, we also considered scenarios with catch-up (CU) vaccination to age 15 or 20 when vaccination was introduced. An overview of vaccination scenarios is shown in Online Supplement Table A.14.

### Model Outcomes

We provide model-based projections of cervical cancer incidence reductions (Online Supplement Table A.15) and disability adjusted life years (DALYs) averted compared to no vaccination, as well as the costs of vaccination and savings from cancer treatment costs. We used these outcomes to compute the incremental costs and health effects relative to no vaccination and return on investment (ROI) of single-dose vaccination. ROI was calculated as the ratio between saved cancer treatment costs and vaccination costs and converted to a percentage. We estimated the incremental cost-effectiveness ratio (ICER) of single-dose versus no vaccination, and of two-dose versus single-dose and no vaccination, using a healthcare payer perspective. ICER was estimated for the whole country and for each Indian state separately.

Model outcomes are presented by year since the start of vaccination. Time horizon of the analysis is 100 years starting at the year in which vaccination was introduced. Costs are shown in $USD (and as a sensitivity analysis in $IUSD, for international comparisons), using 2020 prices and 2020 INR/USD and purchasing power parity (PPP) conversion rates. We used a discount rate of 3% for DALYs and costs as recommended by WHO and also show undiscounted results. Disability weights and durations associated with each cervical cancer health state were based on the Global Burden of Disease 2017 version (Online Supplement Table A.16). We considered two cost-effectiveness thresholds for the ICER, a) 100% of Indian GDP per capita, as per WHO recommendation ($1995), and b) 30% of Indian GDP per capita ($598), based on previous estimates of actual cost-effectiveness thresholds for LMICs.^12-14^

This study adheres to HPV-FRAME, a quality framework for mathematical modelling evaluations of HPV-related cancer control.^27^ The checklist is reported in Online Supplement Tables A.17-A.20.

### Univariate Sensitivity Analyses and Probabilistic Analysis

We varied several economic parameters used for the base-case vaccination scenario. We considered an increase from 3% to 6% in discount rate, a change in cancer treatment costs of ±50%, a change of ±50% in the costs of implementation and delivery of vaccination, and 2% yearly reduction in vaccine price, informed by longitudinal GAVI data on pneumococcal vaccine prices (S Appendix).

In the probabilistic analysis, we included uncertainty distributions for the type-specific duration from CIN2/3 to cancer, 5-year stage-specific cancer survival probabilities, costs of cancer treatment, vaccination implementation and delivery costs. We also included variation in type-specific HPV incidence, based on the 100 best-fitting parameter sets of the transmission model. We ran 100 Monte Carlo simulations of the no vaccination and base-case scenarios, with sampled parameter values based on the distributions shown in Online Supplement Table A.21. Results are shown in a scatterplot and as a cost-acceptability curve. For the base case scenario, we also report an uncertainty interval (UI) based on the 10^th^ and 90^th^ percentiles of the model outcomes.

## RESULTS

### Base Case Scenario

The base-case scenario consisted of single-dose HPV vaccination of girls aged 10 with 90% coverage and lifetime protection (Assumption A). Thirty years after the start of vaccination, annual cancer incidence was projected to decrease by 10% (UI: 4%, 14%). After 50, 75 and 100 years since start of vaccination, annual cancer incidence was projected to decrease by 45% (UI: 39%, 55%), 68% (UI: 63%, 76%) and 72% (UI: 65%, 79%), respectively (Figure 1, Online Supplement Figures B.1-B.6).

**Figure 1:**
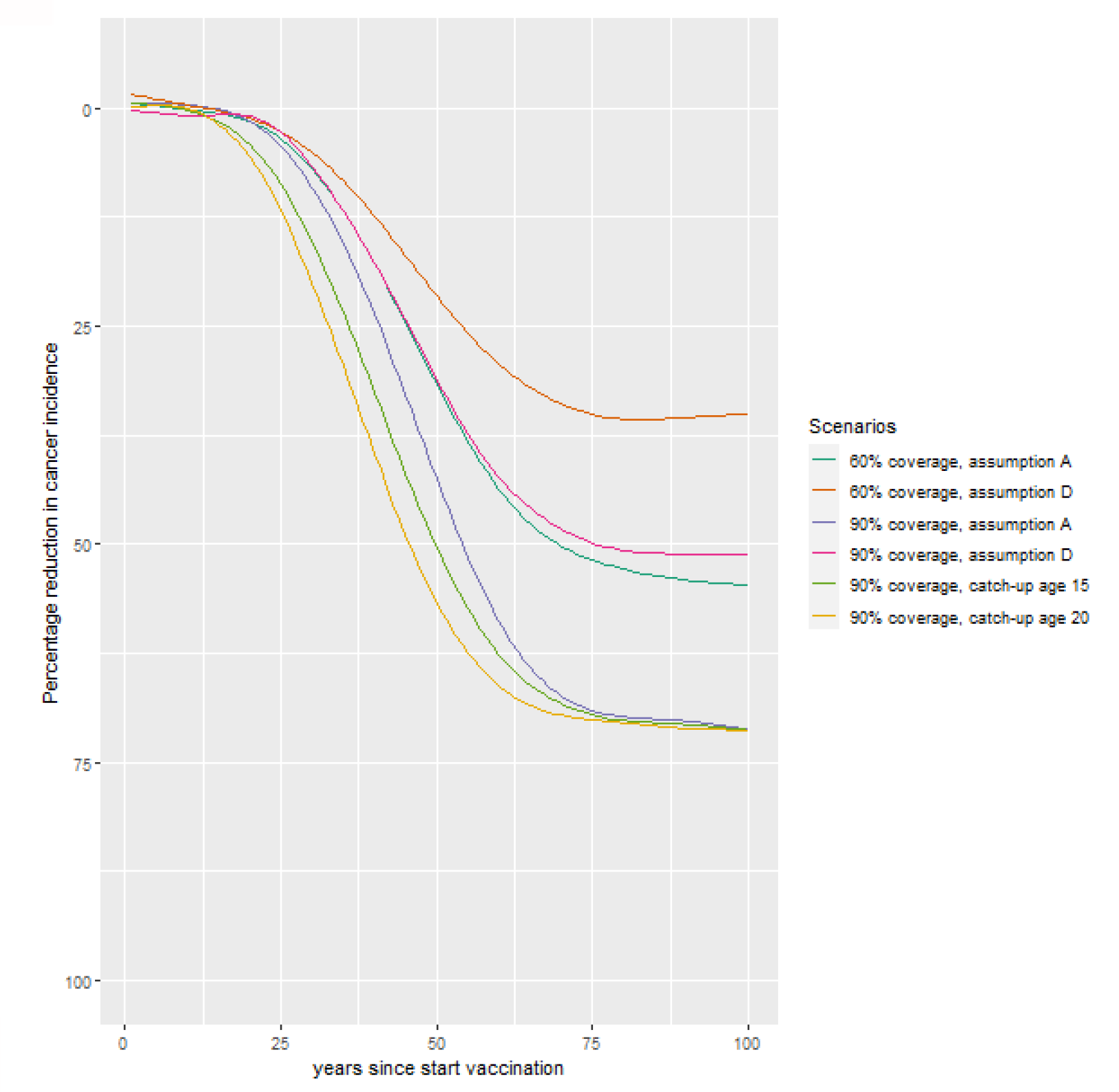
Projected reduction in cervical cancer incidence after introduction of single-dose HPV vaccination in India by scenario.^a^. ^a^ Vaccine protection assumption A denotes a vaccine efficacy of 95% for HPV 16/18, 9% for HPV 31/33/45, and 0% for the remaining oncogenic HPV types, with lifetime protection. Vaccine protection assumption D denotes 85% vaccine efficacy against HPV16, 55% vaccine efficacy agains t HPV18, with exponentially decreasing efficacy during the entire lifetime and remaining efficacy 20 years post-vaccination of 65%. Cross-protection for types HPV 31/33/45 starts at 9% with efficacy waning at the same rate as for HPV18. Catch-up vaccination until age 15 or 20 is shown for 90% coverage in the catch-up cohorts, with vaccine protection assumption A as in the base-case. HPV=human papillomavirus.

The total cost of introducing single-dose HPV vaccination was estimated at about $USD 106 million (UI: $100 million, $112 million) in the first year. This is comparable with 9% of the annual cost of the Indian universal childhood vaccination programme. With no discounting, the intervention would be cost-saving with incremental costs of- $388 thousand per 100,000 women (UI: -$1,607 thousand, -$34 thousand) and a ROI of 32% (UI: 3%, 133%) (Online Supplement Figure B.7). However, when discounting costs and DALYs averted at 3% per year, the costs of vaccination outweighed the costs saved from cancer treatment, resulting in an incremental cost of $167 thousand per 100,000 women and a negative ROI of-42% (UI: - 57%, 10%) (Figure 2). The ICER in the base-case scenario relative to no vaccination was $405 per DALY averted (UI:-$41, $771), which is below the threshold of 30% of the Indian GDP per capita (Figure 2).

**Figure 2:**
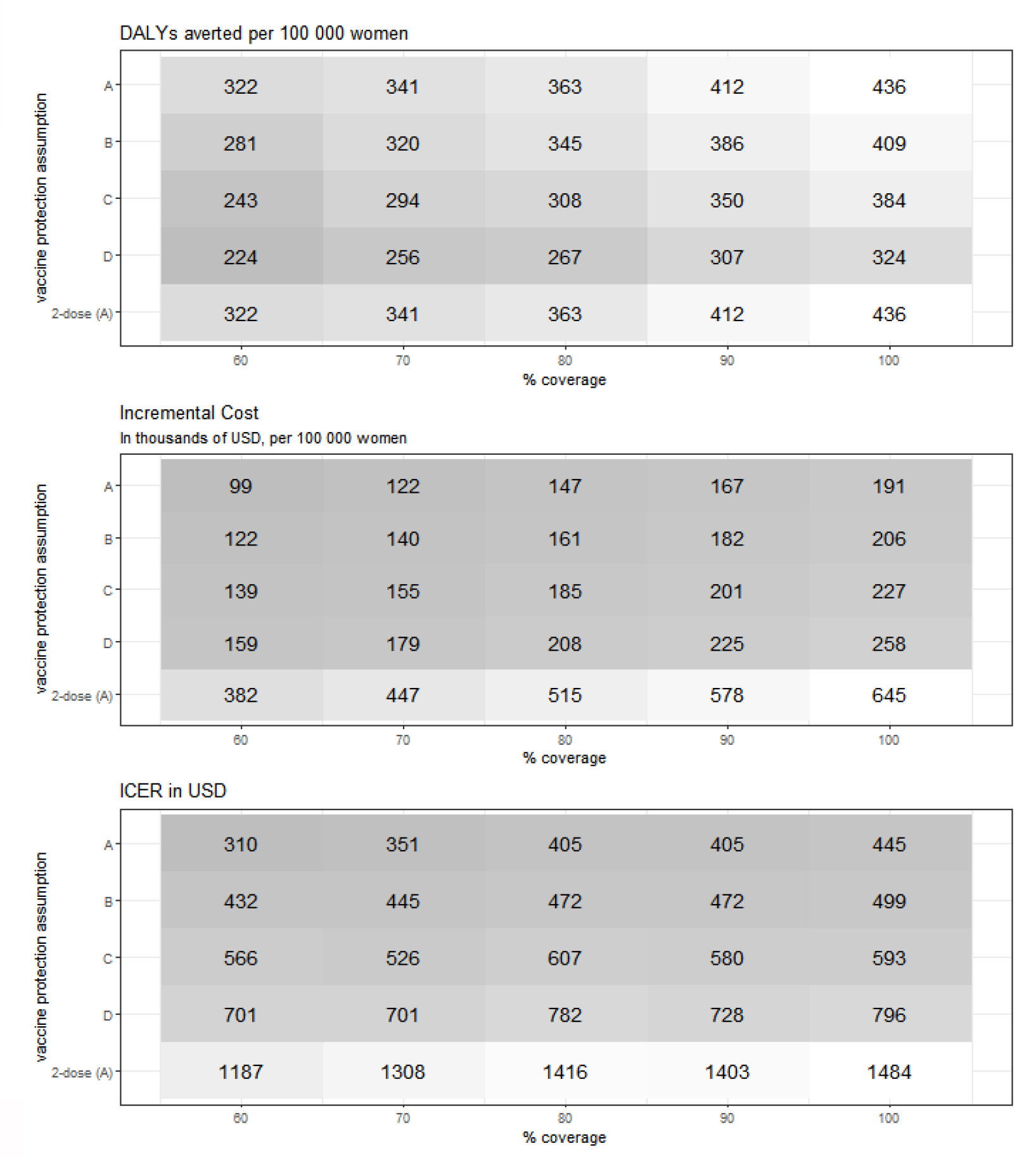
Incremental costs and health effects of single- and two-dose vaccination versus no vaccination.^a^. ^a^ Costs are given in $USD at 2020 prices. Costs and disability-adjusted life years (DALYs) averted are discounted at 3%. ICER denotes incremental cost-effectiveness ratio per DALY averted relative to no vaccination. Darker colours denote lower values.

State-level ICERs are shown in Figure 3 and appendix p. 38. These ranged from $67 to $593 per DALY averted and therefore all states had an ICER below the 30% of GDP per capita threshold. For states classified as high cancer incidence, the ICERs ranged between $67 to $336, while for states classified as low cancer incidence, the ICERs ranged between $220 to $593 per DALY averted.

**Figure 3:**
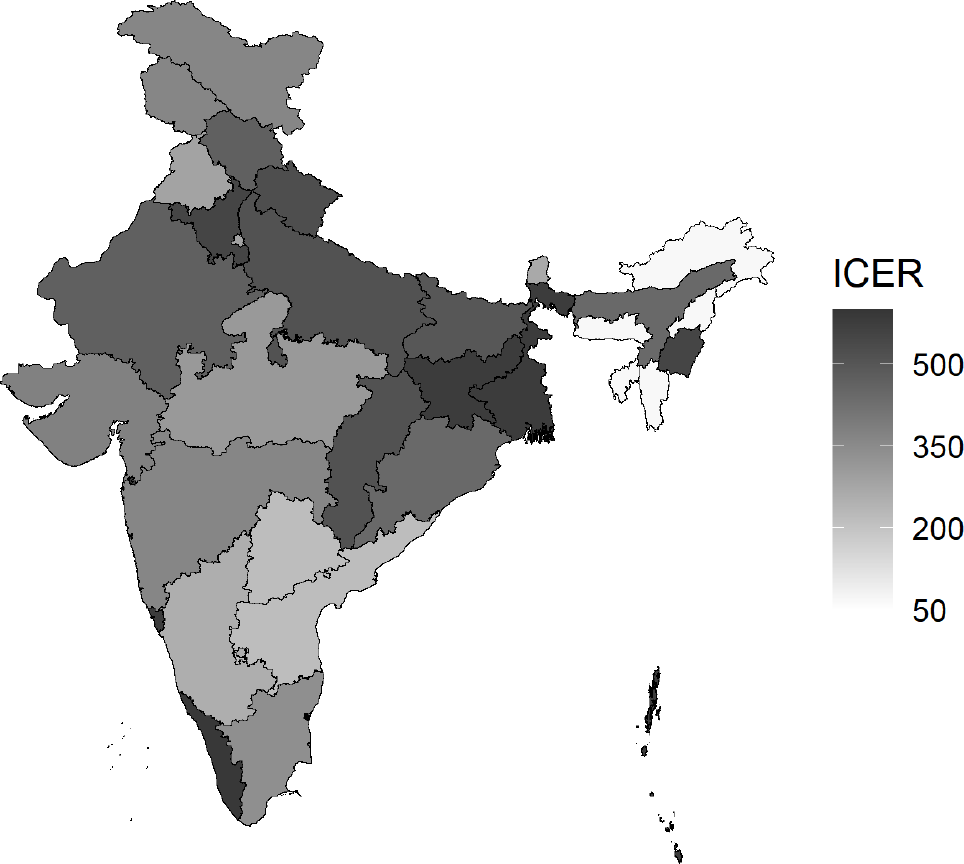
Incremental cost-effectiveness ratio (in $USD) by Indian state for the base case scenario.^a^. ^a^ Base-case scenario denotes single-dose vaccination with 90% uptake among 10-year-old girls, a 95% vaccine efficacy against HPV16/18 with no waning, and 9% cross-protection for types HPV 31/33/45. Costs are given in thousands of $USD at 2020 prices. Costs and disability-adjusted life years (DALYs) averted are discounted at 3%. ICER denotes incremental cost-effectiveness ratio per DALY averted relative to no vaccination.

### Scenarios of single-dose vaccine protection and vaccination coverage

For the worst-case assumption D with 90% coverage, annual cancer incidence decreased by 51% after 100 years since start of vaccination, compared to 72% in the base case. ICERs under 90% coverage (relative to no vaccination) of assumptions B, C, and D were equal to $472, $580 and $728 per DALY averted, respectively, compared to $405 per DALY averted in the base-case. For 60% coverage, the ICER was $310 for assumption A and $701 for assumption D (Figure 2). All scenarios had an ICER below the WHO threshold and all scenarios except D had an ICER below 30% of the Indian GDP per capita (Figure 2).

### Two-dose Vaccination

We assumed that the health effects of two-dose vaccination were the same as for single-dose vaccination in the base case (Assumption A). The total costs of two-dose vaccination in the first year were $200 million. Relative to no vaccination, the incremental costs were $578 thousand per 100,000 women, compared to $167 thousand for single-dose vaccination, and the resulting ICER was $1403 per DALY averted (Figure 2 and Online Supplement Figure B.8). This is about 70% of the Indian GDP per capita. The ICER of two-dose vaccination compared to single-dose vaccination for scenarios under assumptions B, C and D was between $2279 and $19504 per DALY averted (Figure 4), above the WHO threshold ($1995).

**Figure 4:**
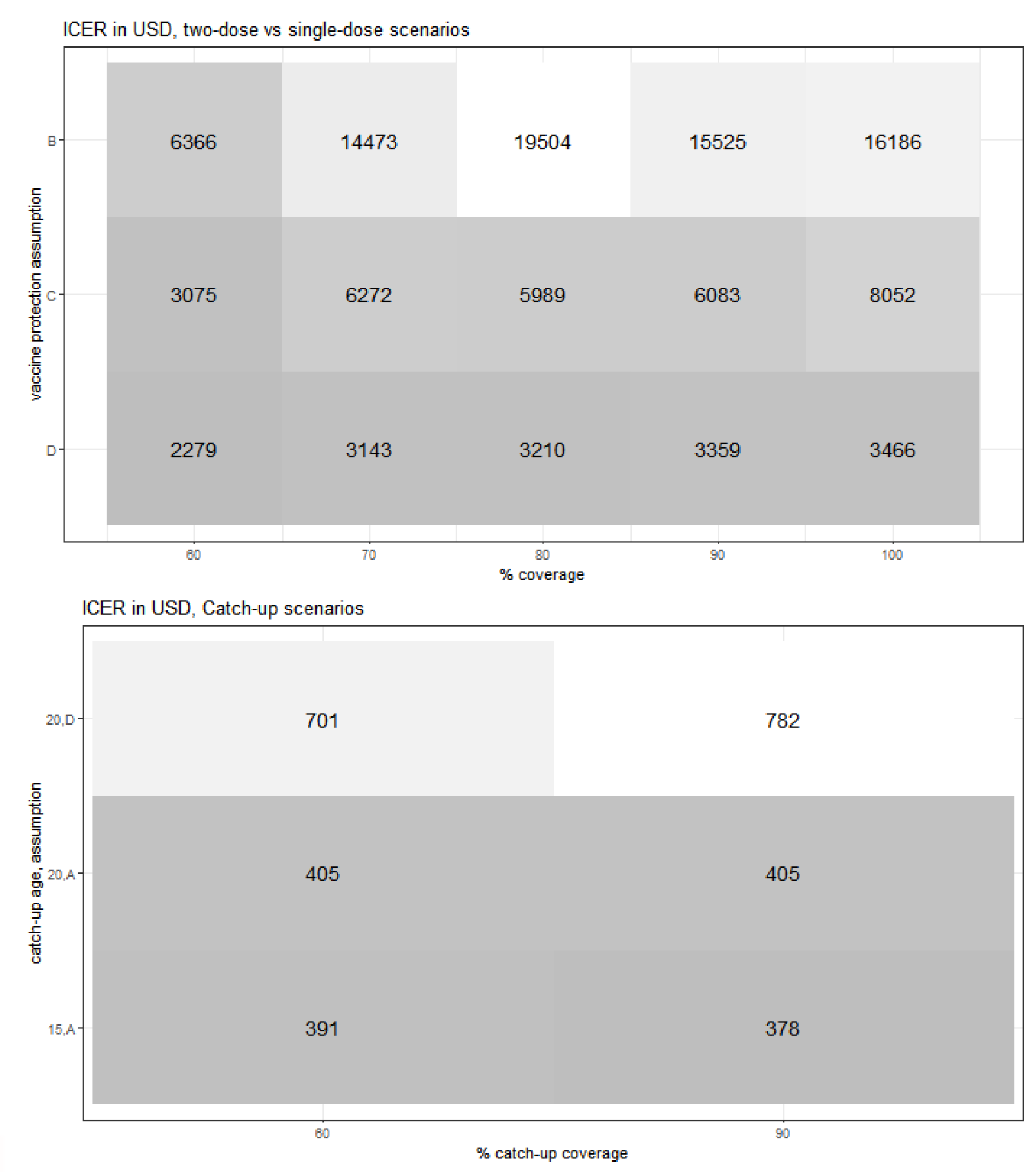
Incremental cost-effectiveness ratio for two-dose versus single-dose vaccination (assuming lifetime protection for two-dose) and for single-dose vaccination with catch-up.^a^. ^a^ Costs and disability-adjusted life years (DALY s) averted are discounted at 3%. Costs are given in $USD at 2020 prices. ICER denotes incremental cost-effectiveness ratio per DALY averted of two-dose vs single-dose (top panel) and relative to no vaccination (bottom panel). In the top panel, Scenario A is not applicable since incremental DALYs of two-dose vaccination is equal to zero. Darker colours denote lower values.

### Catch-Up Vaccination

With catch-up to ages 15 and 20, we predicted a faster decline in annual cervical cancer incidence, for instance, of 16% and 21% after 30 years, instead of 10% without catch-up. Over time, however, annual cervical cancer incidence will become similar to the scenarios without catch-up. The total cost in the first year for catch-up to age 15 was $475 million, which corresponds to 40% of the annual cost of the Indian universal childhood vaccination programme. With catch-up to ages 15 and 20, the ICER ranged between $378 and $405 per DALY averted (Figure 4, Online Supplement Figure B.9).

### Sensitivity Analyses

Valuing costs in $IUSD instead of $USD resulted in ICERs more favourable towards vaccination (Online Supplement Figures B.10-B14). In the cost sensitivity analyses, we found that a 6% discount rate would substantially increase the ICER from $405 in the base-case scenario to $2513 per DALY averted (Figure 5). Decreasing the treatment costs by 50%, or increasing implementation or delivery costs by 50%, resulted in an increase in the ICER to

**Figure 5:**
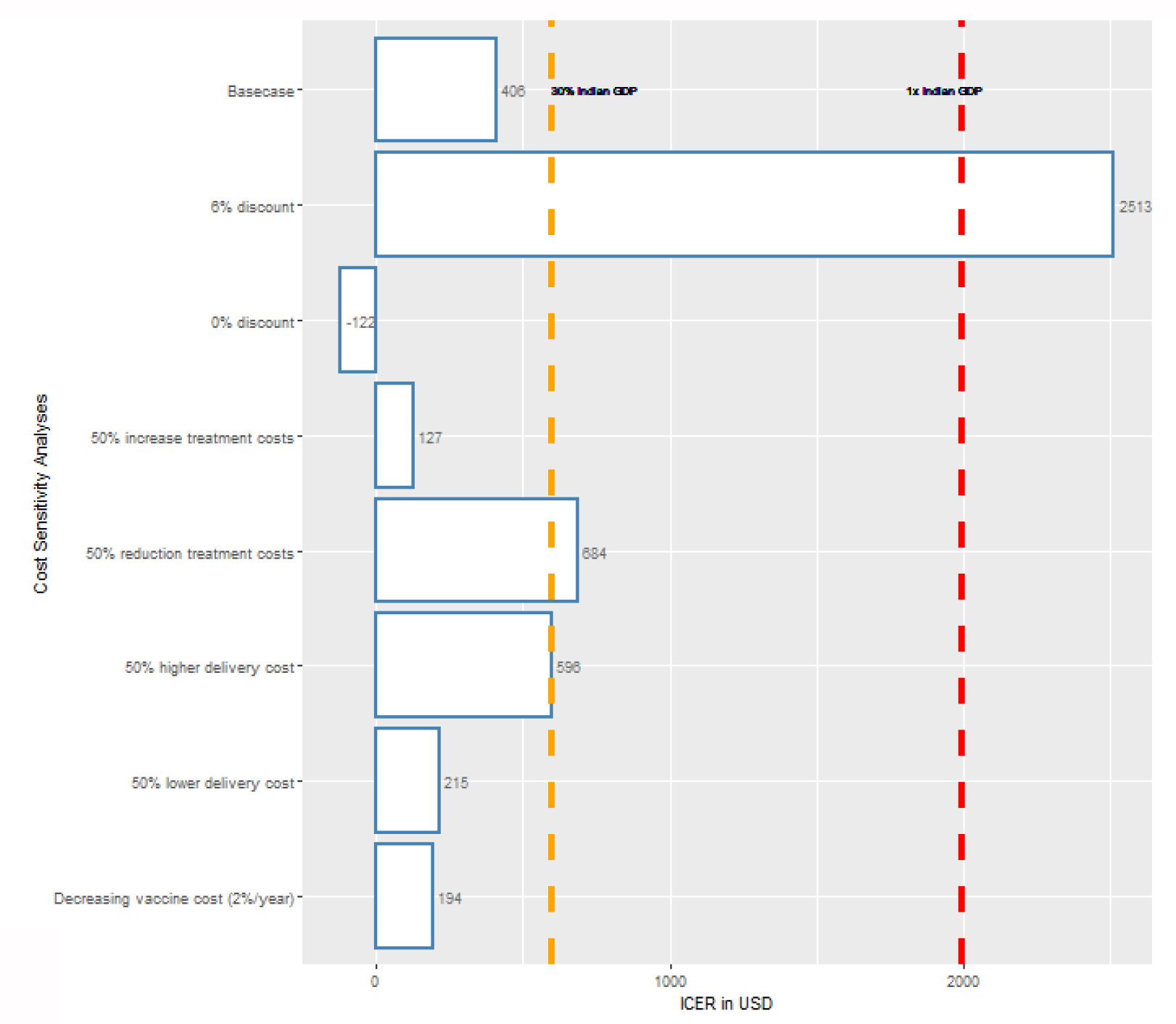
Univariate sensitivity analysis on cost variables. ^a^ Costs are given in $USD at 2020 prices. Costs and disability-adjusted life years (DALYs) averted are discounted at 3% per year in the base case. ICER denotes incremental cost-effectiveness ratio. The dashed lines denote the range for cost-effectiveness thresholds, 30% of Indian GDP per capita in $USD (orange) and 100% of Indian GDP per capita in USD (red). GDP=gross domestic product.

$684 or 596 per DALY averted, respectively. When we assumed that vaccine price would drop at a rate of 2% per year, the ICER would decrease to $127 per DALY averted.

Taking uncertainty in model parameters and cost variables into account via probabilistic analysis, we estimated that single-dose HPV vaccination had approximately 77% probability of being cost-effective at a threshold of 30% of the Indian GDP per capita. Only 1% of the sampled ICERs were above the WHO threshold. By contrast, in 13% of the draws, single-dose vaccination was cost-saving (Figure 6).

**Figure 6:**
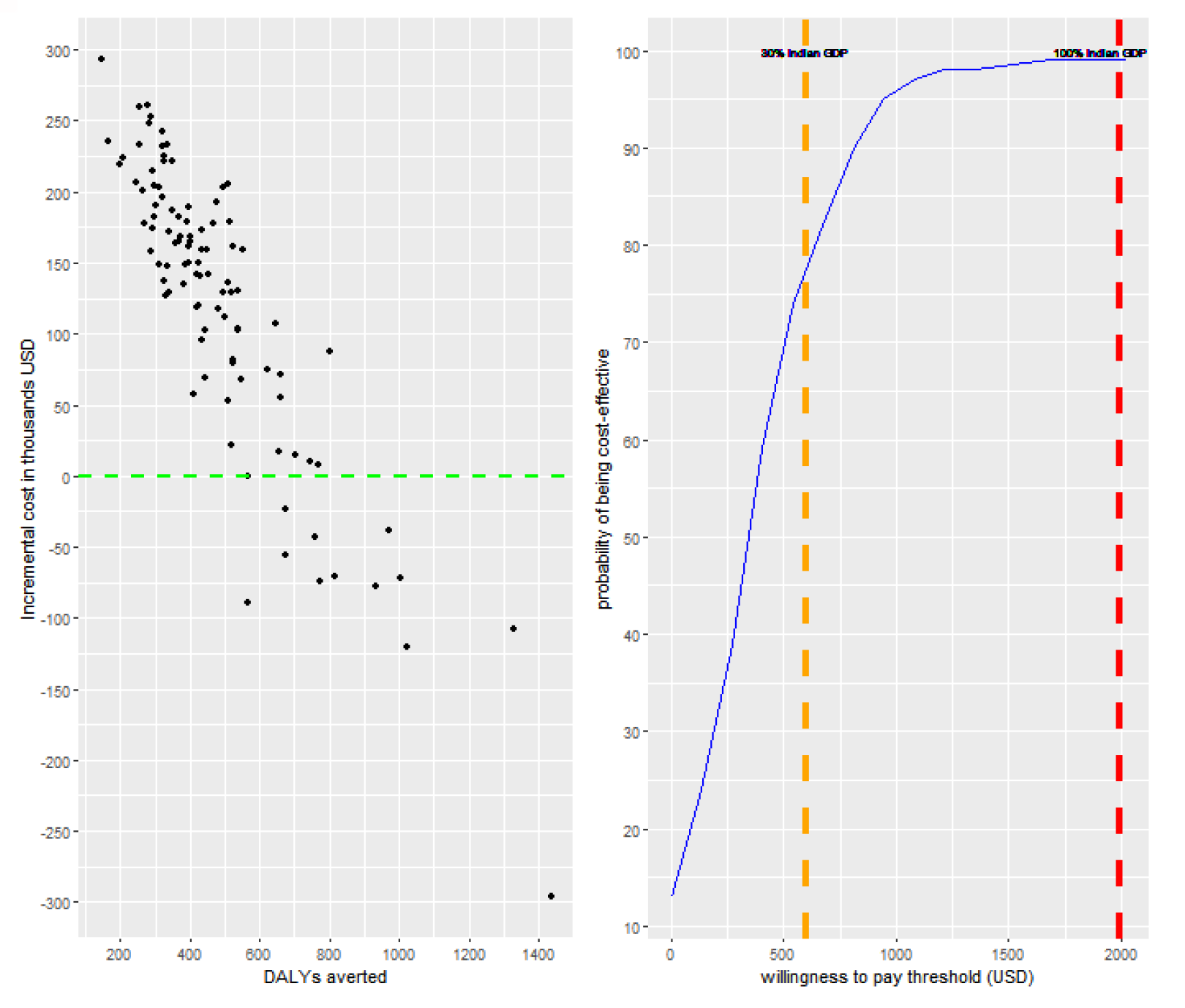
Probabilistic analysis for the base-case scenario of 90% coverage and lifetime protection. ^a^. ^a^ The green dashed line in the left panel denote s the range below which single-dose vaccination is cost-saving. The dashed lines in the right panel denote the range for cost-effectiveness thresholds, 30% of Indian GDP per capita in $USD (orange) and 100% of Indian GDP per capita in $USD (red). GDP=gross domestic product.

## DISCUSSION

HPV vaccination is a key component of the WHO strategy towards elimination of cervical cancer as a public health problem globally. In Man et al. ^16^, we showed that a national immunisation programme relying on single-dose vaccination for 10-year-old girls would be sufficient to meet the WHO-defined elimination target across India. Here, we show that such a programme would have an ICER of $405 per DALY averted, far below the WHO - recommended threshold for cost-effective interventions of 100% GDP per capita ($1995) and a high probability of achieving an ICER below the more realistic willingness-to-pay threshold of 30% GDP per capita, suggested in previous studies. ^12-14^ This corresponds to a total first-year budget impact equivalent to 9% of the annual budget of the Indian universal childhood immunisation programme.^26^ We also show that catch-up single-dose vaccination up to ages 15 or 20 is a cost-effective strategy.

Furthermore, while we found substantial state-level heterogeneity in the ICERs, depending on the projected level of cervical cancer incidence and delivery costs, the 30% threshold was met in every state without exception. On the other hand, we found two-dose HPV vaccination to have a much less favourable cost-effectiveness profile, with an ICER against no vaccination of about 70% of Indian GDP per capita at the current GAVI supported price, and, even for the worst case scenario of single dose protection, an ICER against single-dose vaccination of $2279 per DALY averted, which is well above the WHO threshold ($1995).

The health and economic effects of introducing HPV vaccination in India have been investigated before. Two previous studies have assessed the cost-effectiveness of two-dose vaccination in India,^28,29^ and two studies assessed the cost-effectiveness of single-dose vaccination.^30,31^ All studies valued costs in IUSD and found that HPV vaccination would be cost-effective at a threshold of 100% GDP per capita. While there is no official cost-effectiveness threshold for India, and current willingness-to-pay estimates are uncertain, there is an agreement that a threshold well below 100% of national GDP better reflects opportunity costs for LMICs than the WHO recommendation.^12-14^ Compared to previous studies, our projections are therefore more conservative.

Another strength of our approach, relative to previous studies, is our reliance on context-specific data. Assumptions concerning efficacy and long-term protection given by single-dose HPV vaccination were based on efficacy and immunogenicity data from the IARC India vaccine trial.^9^ We used state-specific data on cervical cancer incidence and sexual behaviour to project the state-specific impact of HPV vaccination. Treatment costs were collected from Indian hospitals and from the literature. Implementation and delivery costs of HPV vaccination were collected from Sikkim state government. Extrapolation to obtain national cost data was informed by a recent district level cost analysis of routine childhood immunisation in India.^26^ This also enabled us to quantify the budgetary requirement for HPV vaccine introduction in India in terms of the Indian universal childhood immunisation programme. Estimates of budget impact and return on investment are often lacking in cost-effectiveness studies of HPV vaccine introduction in LMICs, and have not been previously available for India.

Our study also has limitations. Some of these relate to uncertainty around sexual behaviour and cancer incidence trends in India and are discussed in Man et al.^16^ There are also limitations concerning the economic input. While our treatment costs are based on Indian data, it was difficult to obtain a representative sample of costs for the whole country. Our implementation and delivery costs were extrapolated from Sikkim government data, however this state has a relatively small population. Furthermore, we used a payer perspective and therefore we did not take into account costs related to productivity losses.

Implementing cervical cancer screening along with nationwide introduction of HPV vaccination could further speed up reduction in cervical cancer incidence. However, currently in India, there are multiple cultural, financial and logistical barriers restricting access to screening, with only 2% of women screened in the age group 35–50 years.^24^ If single-dose HPV vaccination is implemented, a screening programme could still be worthwhile to align with the WHO strategic targets for cervical cancer elimination in the near future.

In this mathematical modelling study, we have shown that single-dose HPV vaccine introduction in India is likely to be cost-effective under a stringent willingness-to-pay threshold of 30% of the GDP per capita. Two-dose vaccination would have a less favourable cost-effectiveness profile. These results could be used by Indian government health officials in their decision-making on the introduction of HPV vaccination and could convey several lessons for implementation in other LMICs.

## Data Availability

External researchers can make written requests to the IARC for sharing of data regarding the IARC India vaccine trial. Requests will be assessed on a case-by-case basis in consultation with lead and co-investigators. A brief analysis plan and data request will be required and reviewed by the investigators for approval of data sharing. When requests are approved, anonymized data will be sent electronically in password protected files. All data sharing will abide by rules and policies defined by the involved parties. Data sharing mechanisms will ensure that the rights and privacy of individuals participating in research will be protected at all times. Data from public sources are listed in the appendix (pp 7-16). The model code is available from the authors on request.

## CONTRIBUTORS

IB, PBa, and RS contributed to funding acquisition of study. The project was supervised by IB. TMdC, JB, JAB, and IB co-designed and co-led the formal analysis, investigation, validation and visualisation of the study findings. IM and DG contributed to the investigation and visualisation of the analyses and study findings. TMdC, JAB, and JB led the development of modelling software. TMdC, IM, DG, PB, LRS, RM, EL, KI, and SM were responsible for data curation. TMdC and JAB accessed and verified all reported data.

All authors had full access to all of the data reported in the study. TMdC, JB, and JAB drafted the original draft of the manuscript. All authors contributed to reviewing, editing, a nd approved the final manuscript. All authors had final responsibility to submit for publication.

## ACKNOWLEDGMENTS

This study was funded by the Bill & Melinda Gates Foundation (grant numbers: OPP48979; INV-039876). For the authors identified as personnel of the International Agency for Research on Cancer or World Health Organization, the authors alone are responsible for the views expressed in this article and they do not necessarily represent the decisions, policies or views of the International Agency for Research on Cancer or World Health Organization. The designations used and the presentation of the material in this Article do not imply the expression of any opinion whatsoever on the part of WHO and the IARC about the legal status of any country, territory, city, or area, or of its authorities, or concerning the delimitation of its frontiers or boundaries.

