## Supplementary material for "Health and economic effects of introducing single-dose human papillomavirus vaccination in India": Online Supplement

### Technical Appendix for: Health economic impact of introducing single-dose HPV vaccination in India.

**List of figures**

Figure A.1: Natural history from infection to cervical cancer.
Figure A.2:  Observed and predicted cancer incidence per 100 000 women.

**List of tables**

Table A.1. List of model parameters.

Table A.2: Stage distribution observed and predicted by the cancer progression model

Table A.3: Hysterectomy rates by age group, adapted from Meher and Sahoo 2020.

Table A.4: 5-year cervical cancer survival by cancer registry/hospital and data sources.

Table A.5: Currencies and conversion rates.

Table A.6: Initial Cervical Cancer Treatment Costs in INR.

Table A.7: Additional input for treatment costs calculation.

Table A.8: Cervical Cancer Treatment Costs per stage.

Table A.9: Original cost items for communication and delivery costs of Sikkim programme.

Table A.10: Calculations for two-dose and single-dose vaccination costs in Sikkim (2020 prices).

Table A.11: Breakdown of estimated costs per dose for Sikkim.

Table A.12: Extrapolation delivery costs Sikkim to India based on costs of childhood vaccination programme.

Table A.13: Overview of single-dose waning scenario parameters.

Table A.14: Vaccination Scenarios.

Table A.15: Age-specific cervical cancer incidence (per 100 000 women) by Indian state.

Table A.16: Disability weights and durations for different phases of cervical cancer in GBD 2017 study.

Table A.17: HPV-FRAME Checklist, Core Reporting Standard.

Table A.18: HPV-FRAME Checklist, Reporting standard for HPV vaccination in adolescent individuals.

Table A.19: HPV-FRAME Checklist, Reporting standard for models of HPV prevention in LMIC.

Table A.20: HPV-FRAME Checklist, Reporting standards for evaluations assessing alternative vaccine types or reduced-dose schedules.

Table A.21: Sampling distributions for model parameters included in probabilistic analysis.

#### A.1 HPV transmission model and progression model

##### A.1.1 HPV transmission model

The HPV transmission model (EpiMetHeos) is an extension of EpiModel, an open-source statistical framework that allows simulation of infectious disease transmission on dynamic contact networks [1]. This model is extensively described in Man et al [2].

##### A.1.2 Cancer progression model

The cancer progression model is an individual-based discrete-time microsimulation model that simulates birth cohorts of women from age 10 to death, in six-month time steps. The pre-invasive part of the model consists of 13 parallel Markov chains, corresponding to an infection with HPV type 16, 18, 31, 33, 35, 39, 45, 51, 52, 56, 58, 59 and 68. The 6-month cumulative probabilities of type-specific HPV infection for each 6-month age-birth cohort were calculated by the HPV transmission model (EpiMetHeos).

The structure of the cancer progression model with regard to the natural history of type-specific high-risk HPV infection conforms to that of the HPV transmission model, up to CIN2/3 for each high-risk HPV type. The duration of infection follows a type-specific distribution with six parameters $\gamma_i, \eta_i, \delta_{i,1}, \delta_{i,2}, \nu_{i,1}, \nu_{i,2}$ , where $i$ denotes the HPV type. These parameters are shared between the HPV transmission model and the cancer progression model. Some of these parameters were estimated in Bogaards et al. 2010[3] and also used in Bogaards et al. 2011 [4] and Berkhof et al. 2013 [5], namely: i) $\eta_i$ which denote genotype-specific progression rates from state HPV infection (CIN0) to CIN1; ii) $\gamma_i$ and $\delta_{i,1}$ which denote the clearance rates for CIN0 and CIN1 stages. Other parameters, namely, iii) $\nu_{i,1}$ and $\nu_{i,2}$ which denote the rates of progression from CIN1 to CIN2/3 regressive and non-regressive stages and iv) $\delta_{i,2}$ the clearance rate from the regressive CIN2/3 stage were calibrated simultaneously using data from the POBASCAM trial.[6] This was done by comparing simulated model estimates to age-specific frequencies of CIN2/3 cases, cancer cases and the proportion of HPV16 positive and HPV18 positive CIN2/3 and cancer cases. Likelihood was maximised by stochastic optimisation where the exact likelihood is replaced by a simulation run estimate. For this we used the simultaneous perturbation stochastic approximation algorithm.[7] Additionally, for this study, the progression, regression, and loss of immunity parameters of non-HPV16/18 infections were set equal across genotypes by weighted pooling.[5]

The duration between CIN2/3 and cervical cancer was modelled as a gamma distribution (distinct for HPV types 16 and 18 versus other types) and estimated based on Dutch cancer registry data.[8] Progression between invasive cancer states FIGO1a, FIGO1b and FIGO2+ was based on national registry and screening data [5] (Figure A.1, TableA.1).

**Figure A.1: Natural history from infection to cervical cancer.**

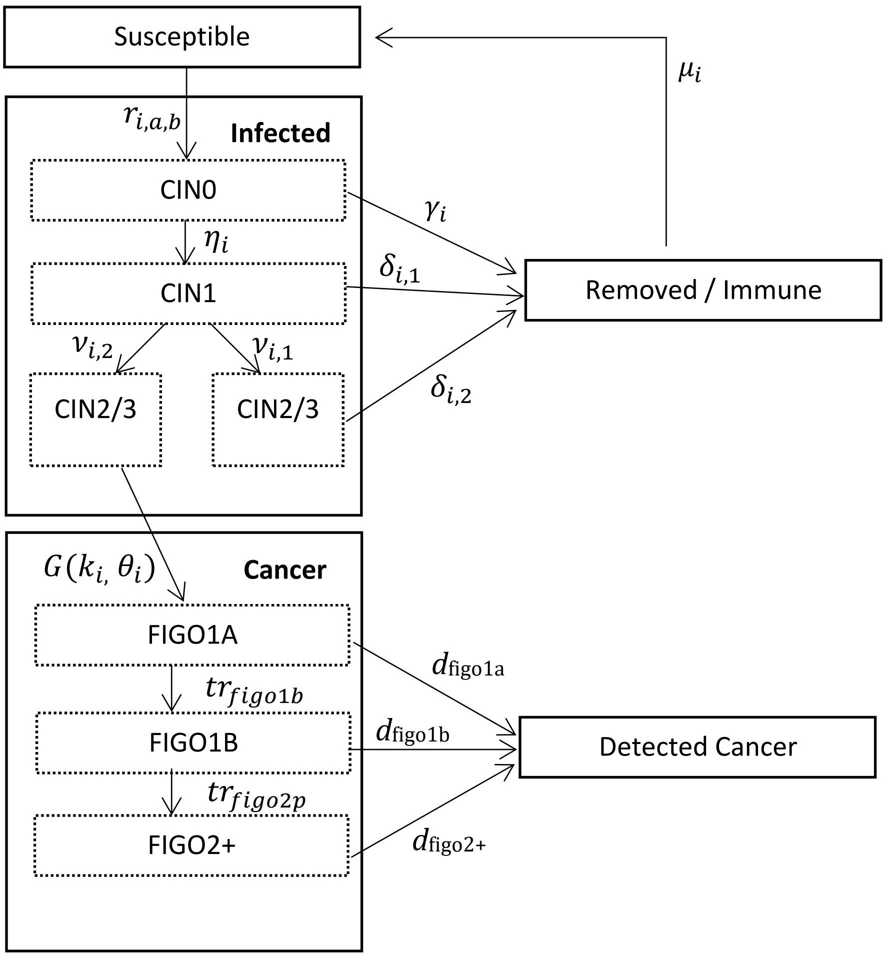

**Table A.1: List of model parameters.**[a]

| Notation | Description | Values | Reference |
|---|---|---|---|
| *Natural History of Infection* | | | |
| $r_{i,a,b}$ | Forces of infection for HPV16, HPV18, cross-protected types and non-cross-protected types. | Range of values per HPV type (*i*) , age group (*a*) and birth cohort (*b*). | Computed using EpiMetheos. |
| $\gamma_{16}, \gamma_{18}, \gamma_{other}$ | Clearance rate from CIN0. | 0·29 ; 0·34 ; 0·41[b] | Berkhof et al 2013 [5] |
| $\eta_{16}, \eta_{18}, \eta_{other}$ | Progression rate from CIN0 to CIN1. | 0·24 ; 0·19 ; 0·11 | Berkhof et al 2013 [5] |
| $\delta_{16,1}, \delta_{18,1}, \delta_{other,1}$ | Clearance rate from CIN1. | 0·06 ; 0·17 ; 0·21 | Berkhof et al 2013 [5] |
| $\delta_{16,2}, \delta_{18,2}, \delta_{other,2}$ | Clearance rate from regressive CIN2/3. | 0·65 ; 0·65 ; 0·65 | Section A.1.2 |
| $\nu_{16,1}, \nu_{18,1}, \nu_{other,1}$ | Progression rate from CIN1 to regressive CIN2/3. | 0·02 ; <0·01 ; 0·02 | Section A.1.2 |
| $\nu_{16,2}, \nu_{18,2}, \nu_{other,2}$ | Progression rate from CIN1 to non-regressive CIN2/3. | 0·02 ; 0.02 ; 0·01 | Section A.1.2 |
| $\mu_{16}, \mu_{18}, \mu_{other}$ | Rate of waning natural immunity. | 0·02 ; 0·01 ; 0·02 | Berkhof et al 2013 [5] |
| *Duration to Cancer* | | | |
| $k_{16}, k_{18}, k_{other}$ | Shape parameter of gamma distribution. | 9·67 ; 9·67; 2·49 [b] | Vink et al 2013 [8] |
| $\theta_{16}, \theta_{18}, \theta_{other}$ | Scale parameter of gamma distribution. | 3·33 ; 3·33 ; 9·14 | Vink et al 2013 [8] |
| *Cancer progression & detection* | | | |
| $d_{figo1a}, d_{figo1b}, d_{figo2p}$ | Detection rates for FIGO1a/FIGO1b/FIGO2+ health states in absence of screening. | 0·0125 ; 0·025 ; 0·3 | FIGO1a, FIGO1b: Calibrated (Table A.2). FIGO2+: Berkhof et al 2013 [5] |
| $tr_{figo1b}, tr_{figo2+}$ | Transition rate from FIGO1a to FIGO1b and from FIGO1b to FIGO2+ | 0·125 ; 0·1 | Berkhof et al 2013 [5] |

[a] All rates per 6-month period, unless otherwise indicated.

[b] Respectively, HPV16, HPV18 and Other HPV strains (31, 33, 35, 39, 45, 51, 52, 56, 58, 59 and 68).

##### A.1.3 Model Calibration

The calibration of the HPV transmission model is extensively described in Man et al.[2] In short, since high quality HPV prevalence data is not available for every Indian state, we first applied a cluster analysis ("footprinting" as explained elsewhere[9]) to classify states into "high" and "low" cancer incidence clusters based on cancer registry data.[10,11] We then selected one representative state of the high cancer incidence pattern (Tamil Nadu) and one of the low cancer incidence pattern (West Bengal), for which high quality HPV prevalence data was available [12,13] to calibrate the HPV transmission model. Finally, the estimates from these two models were used to extrapolate to all other states within the same cancer incidence patterns cluster.

The natural history from HPV infection to cervical cancer is based on calibration to data in the Netherlands and was assumed to be applicable to India conditional on the local type-specific forces of infection which are computed using the HPV transmission model (EpiMetHeos). This approach was used before to adapt the model to several European countries.[5,14] However, we also adjusted the cancer detection rates to match the observed stage distribution in India. This is justified by the fact that, in absence of organised screening, and due to financial and/or logistic constraints, women in India are likely to be diagnosed at a later time compared to women in the Netherlands.

In Figures A.2-A.3 we show the cancer incidence fit for the Tamil Nadu and West Bengal states. For Tamil Nadu there were several time periods available including 2008-2012,[10] 2012-2013,[15] 2012-2016 [11] and 2017.[16] For West Bengal there was only one time period available: 2012-2016.[11] In Table A.2 we show the stage distribution before and after adjustment of detection rates. The adjustment consisted of reducing the detection rates per 6-month time period for FIGO1a and FIGO1b by 50% ($d_{figo1a}$, $d_{figo1b}$) .

**Figure A.2: Observed and predicted cancer incidence per 100 000 women.** [a,b]

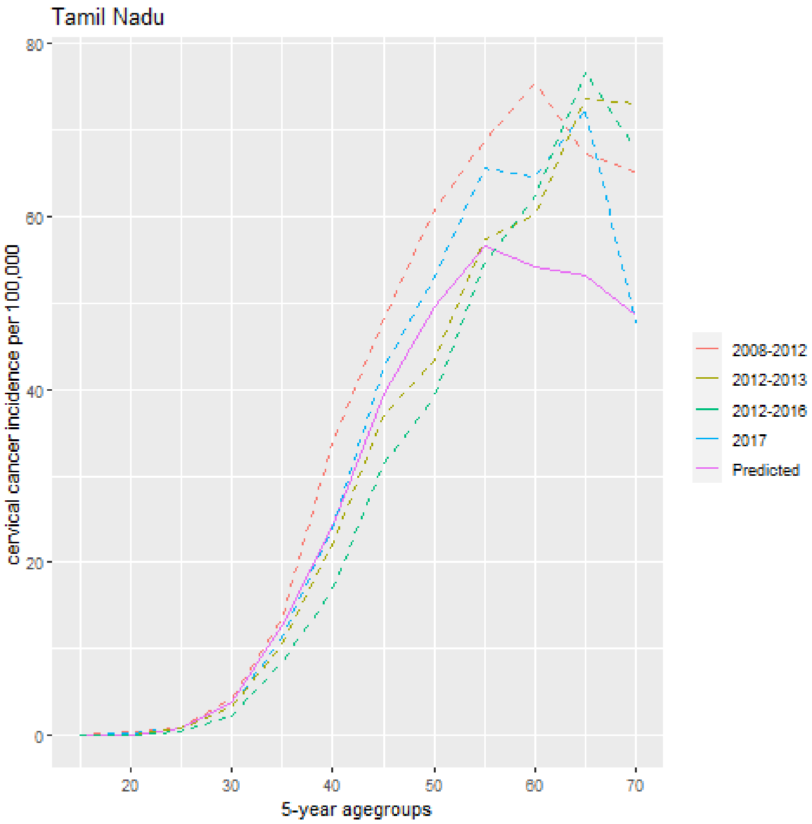

[a] 2008-2012: Average of Chennai and Dindigul registries. 2012-2013, 2012-2016: Chennai registry only, 2017: Tamil Nadu (state, average per district).

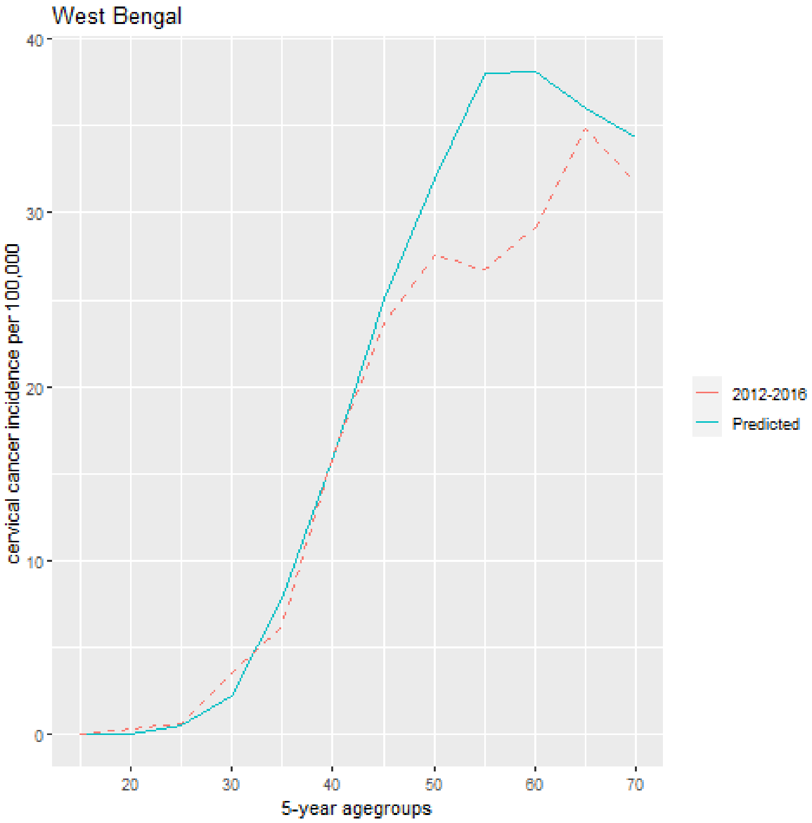

[b] For West Bengal no data from other time periods was available.

**Table A.2: Stage distribution observed and predicted by the cancer progression model.[a]**

| | Localised (FIGO1ab) | Loco regional/Distant (FIGO2+) |
|---|---|---|
| Observed | 33·5% | 66·5% |
| Predicted before adjustment | 45·7% | 54·3% |
| Predicted after adjustment | 28·9% | 71·1% |

[a] Here given the data source (Report of National Cancer Registry Program 2012-2016 [11]), we assumed that localised cancers map 1 to 1 with FIGO1a and FIGO1b health states in the model, and that FIGO2+ denote loco regional and/or distant tumors.

#### A.2 Demographic and Health Parameters

##### A.2.1 Demography

Both the life table and age distribution for women are based on the 2019 version of World Population Prospects UN population projections for India.[17] Life table is based on an estimate for birth cohorts 2005-2010 and age distribution is based on an estimate for 2020.

##### A.2.2 Hysterectomy

Women who undergo hysterectomy are removed from the risk set for cervical cancer. Hysterectomy rates in India are based on Meher and Sahoo 2020.[18] This publication uses data from National Family Health Survey Round 4, a survey including more than 200 000 women. We extrapolated the numbers for all 6-month age groups, by assuming that the observed rates equal the rates in the middle of the age group. For example, 3·7% corresponds to the cumulative probability of hysterectomy at age 34·5 years. We also assume that the hysterectomy rate differs only between age groups 15−29, 30−39 and 40−49 years and that the hysterectomy rate beyond age 50 is similar to the rate in the age group 15−29 years.

**Table A.3: Hysterectomy rates by age group, adapted from Meher and Sahoo 2020. [18]**

| Age Group (years) | Percentage of women with hysterectomy | Sample Size |
|---|---|---|
| 15−29 | 0·6% | 205 603 |
| 30−39 | 3·7% | 184 077 |
| 40−49 | 9·3% | 150 991 |

##### A.2.3 Cancer Survival

Cancer survival is assigned at the time of diagnosis. Since stage-specific survival data for India is not available, we assign the same survival for every cancer stage. We calculated the five-year cancer survival by taking the weighted average of the SurvCan II registries,[19] augmented with newer publications concerning cervical cancer cases diagnosed after the year 2000. For this we performed a literature search on PubMed using keywords "cervical cancer survival India". Out of 890 search results, only 8 results were relevant to our search. Of these, 2 studies were excluded: a) Balasubramaniam et al 2020 [20] was excluded to avoid possible double counting with Nandakumar et al 2015 [21] (since Balasubramaniam et al 2020 [20] uses data from one hospital which is included in Nandakumar et al 2015) ; b) Krishnatreya et al 2016 [22] was excluded since endpoint is 50 month survival and therefore it does not report 5-year survival.

**Table A.4: 5-year cervical cancer survival by cancer registry/hospital and data sources.**

| Cancer Registry/Hospital | N | 5-year Survival | Time of Diagnosis | Source |
|---|---|---|---|---|
| Barshi | 406 | 0·351 | 1990−2000 | SurvcanII [19] |
| Bhopal | 332 | 0·354 | 1990−2001 | SurvcanII |
| Chennai | 4438 | 0·594 | 1990−2000 | SurvcanII |
| Mumbai | 4436 | 0·461 | 1990−2000 | SurvcanII |
| Maharashtra | 192 | 0·432 | 2000−2013 | Jayant et al [23] |
| Dindigul | 223 | 0·350 | 2003−2006 | Swaminathan et al 2009 [24] |
| 12 Indian Hospitals | 2562 | 0·620 | 2006−2008 | Nandakumar et al 2015 [21] |
| Guwahati | 193 | 0·407 | 2010 | Kataki et al 2018 [25] |
| Malabar Cancer Center, Kerala | 227 | 0·668 | 2010−2011 | Bindu et al 2017 [26] |
| Karunagapally | 338 | 0·590 | 2010−2014 | CONCORD3 [27] |
| Trivandrum | 425 | 0·534 | 2012−2014 | Matthew et al 2020 [28] |
| *All* | *19 196* | *0·558* | | |

#### A.3. Economic Parameters

##### A.3.1 Currencies and conversions

We value all costs in 2020 prices. Costs estimates from previous years are converted to 2020 prices using consumer price index for India as estimated by the World Bank.[29] In the base case we value all costs in 2020 $USD. We use an exchange rate (INR per U.S. Dollar) of 74.1. We also value costs in IUSD, a currency which facilitates comparisons between different countries taking into account differences in purchasing power. We use a purchasing power parity exchange rate (Local Currency Units per International Dollar) of 22 [30]. In order to value costs in IUSD, we use the USD/INR exchange rate for tradable goods (e.g. vaccine dose cost), since the price of tradable goods is independent of the country setting and therefore in this case 1 USD = 1 IUSD. For non-tradable goods (e.g. costs medical staff), we use the purchasing power parity exchange rate. For more details on currency conversions to IUSD see the online supplement of Diaz et al 2008. [31]

**Table A.5: Currencies and conversion rates.**

| Conversion | Conversion Rate | Source |
|---|---|---|
| USD/INR | 74·1 | World Bank [29] |
| IUSD/INR | 74·1 (tradable goods)<br>22 (non-tradable goods) | World Bank<br>OECD [30] |

##### A.3.2 Cervical cancer treatment costs

*Overview*

We consider three disease stages for treatment costs, FIGO1a, FIGO1b and FIGO2+ and two types of treatment: 1) radical hysterectomy (RH), which we assume to be performed only to FIGO1a patients, and 2) chemo-radiotherapy (CRT), which is performed to patients with FIGO1b or FIGO2+ at diagnosis. Additionally, we consider rates of recurrence for FIGO1a, FIGO1b and FIGO2+ and we assume that in case of recurrence the patient is assigned to an additional round of treatment and corresponding costs. We distinguish between two types of hospitals, public and private, with different costs per treatment. Both the price differential between public and private hospitals and the proportion of public and private hospital users in the population is based on 75th National Sample Survey data [32] (Tables A.6-A.8).

*Data Collection & Literature Search*

Data from two hospitals (Delhi Surgical Centre of India International Hospital, Regional Cancer Center Trivandrum) was collected. Data from Tata Memorial Center was accessible online.[33] On February 2022, we performed a literature search on PubMed using the following keywords “cervical cancer treatment cost India”. The search returned 65 results. We scanned each result’s title and abstract to verify whether the article was related to cervical cancer treatment costs. Only one publication, Singh et al 2020 [34] (“Cost of Treatment for Cervical Cancer in India “) was related to cervical cancer treatment costs.

*Calculation Steps*

- Step 1: We set all costs to 2020 prices, which affected the costs extracted from the publication, which were collected in 2017. These costs are shown in Table A.6.
- Step 2: We multiply the cost by 4·1 (based on 75th NSS round data [32]) to obtain the costs for a private hospital and calculate the cost as a weighted average between public and private hospital with proportion of private hospital users based on NSS data (Table A.7).
- Step 3: We multiply the average costs with the rate of recurrence per stage, based on data from the literature. The end result is shown in Table A.8.

**Table A.6: Initial Cervical Cancer Treatment Costs in INR.**

| Hospital | Radical Hysterectomy | Chemoradiotherapy |
|---|---|---|
| Delhi SCI | 125 000 | 40 000 |
| Tata Memorial | 64 575 | 90 405 |
| Trivandrum | 67 000 | 89 067 |
| Singh et al 2020 [34] (not reported) | 64 994 | 69 459 |

**Table A.7: Additional input for treatment costs calculation.**

| Description | Value | Source |
|---|---|---|
| Cost multiplier private/public hospitals | 4·1 | 75th NSS round [32] |
| Proportion users private hospitals [a] | 57% | 75th NSS round |
| Recurrence Rate FIGO 1a | 1·5% | Taarnhoj et al 2017 [35] |
| Recurrence Rate FIGO 1b | 7·9% | Uppal et al 2019 [36] |
| Recurrence rate FIGO2+ | 38% | De Foucher et al 2019 [37] |

[a] We do not consider charity hospitals as they represent a proportion of less than 5% of treated women.

**Table A.8: Cervical Cancer Treatment Costs per stage.**

| Average Cost per Step | Radical Hysterectomy (FIGO1a) | Chemoradiotherapy (FIGO1b) | Chemoradiotherapy (FIGO2+) |
|---|---|---|---|
| Public Hospitals | 80 392 | 72 233 | 72 233 |
| Private Hospitals | 333 082 | 299 275 | 299 275 |
| Weighted average Public/Private | 224 007 | 201 271 | 201 271 |
| Total (with recurrence) | 227 307 | 217 193 | 277 754 |

##### A.3.3 Costs of Vaccination

*Overview*

The cost of the vaccine is set equal to the GAVI price of $4·5 USD. The delivery costs were extracted from a government pilot programme in Sikkim. In order to obtain a nationwide estimate for delivery costs in India, we extrapolated these, based on estimated delivery costs for the universal childhood vaccination programme.[38] For this, we assumed that the relative difference in delivery costs per capita between Sikkim and all other states would be the same for childhood vaccination and HPV vaccination.

*Costs Supplied by the Sikkim Government*

The total costs of the two-dose HPV vaccination programme for 2018-2019 were supplied by Sikkim government and are shown in Table A.9. The total number of target girls was calculated as 41 351. This is based on the census 2011 count (38 975) and projected population growth between 2011 and 2018 of 6%.[39] For the conversion from INR to IUSD, we considered vaccine, syringe and hub cutter to be tradable goods. All other cost categories were considered non-tradable.

**Table A.9: Original cost items for communication and delivery costs of Sikkim program.**

| Cost Item | INR |
|---|---|
| Vaccine Costs[a] | 20 987 688 |
| Program Launch | 300 000 |
| Vaccine transportation | 250 000 |
| Syringe and hub cutter [b] | 555 000 |
| Information and communication | 1 850 000 |
| Training | 186 750 |
| Cold Chain | 498 658 |
| Mobility Support for District and state level monitors | 520 000 |
| State overhead costs (14%) | 3 464 733 |
| District Level Costs | 11 006 750 |
| Total Costs | 39 219 579 |

[a] 71 500 doses at GAVI price.
[b] assuming 65 000 syringes and 400 hub cutters.

These costs occurred in 2018/2019 and had to be converted to 2020 prices. We also had to make some adjustments to the total cost, to obtain an estimate for single-dose delivery costs. These adjustments are described in Table A.10. In Table A.11 a breakdown of the costs per target girl for Sikkim is given, and in Table A.12 we show the extrapolation of the costs for the rest of India.

**Table A.10: Calculations for two-dose and single-dose vaccination costs in Sikkim (2020 prices).**

| Step | Description | INR | Explanation |
|---|---|---|---|
| 1 | Total Costs | 39 219 579 | Raw total costs |
| 2 | Use 2020 INR/USD conversion rate for vaccine costs | 42 170 770 | Take depreciation of INR into account |
| 3 | Inflate non-vaccine costs from 2019 to 2020 prices | 43 375 346 | Using Indian CPI rate 1.07 (World Bank) |
| | Total Costs 2-dose 1st Year | 43 375 346 | |
| 4 | Total Costs 2-dose 2nd Year and later | 39 693 258 | Removal of cold chain and programme launch cost, miscellaneous cold chain cost divided by 4 (every 4 years cost, e.g. cold chain maintenance), communication cost and training cost divided by 4. State overhead and district-level costs decrease proportionally to the reduction in costs in year > 1. |
| 5 | Total Costs single-dose 1st Year | 22 985 830 | Removal of remove 50% vaccine costs, state overhead costs (these are proportional to the total cost), syringe costs, mobility support state-level monitors (this cost is by vaccination round), transportation costs and district level costs |
| 6 | Total Costs single-dose 2nd Year and later | 19 626 918 | Removal of cold chain and programme launch cost, miscellaneous cold chain cost divided by 4 (every 4 years cost, e.g. cold chain maintenance), communication cost and training cost divided by 4. State overhead and district-level costs decrease proportionally to the reduction in costs in year > 1. |

**Table A.11: Breakdown of estimated costs per dose for Sikkim.**

| | INR | USD |
|---|---|---|
| Two-dose costs | | |
| Vaccine Cost | 667 | 9·0 |
| Delivery/Programme Cost (Year 1) | 330 | 4·4 |
| Delivery Cost (Year>1) | 293 | 3·9 |
| Communication Cost (Year 1) | 52 | 0·7 |
| Communication Cost (Year>1) | 11 | 0·2 |
| two-dose cost (Year 1) | 1049 | 14·1 |
| Two-dose cost (Year >1) | 960 | 12·9 |
| Single Dose Costs | | |
| Vaccine Cost | 334 | 4·5 |
| Delivery/Programme Cost (Year 1) | 170 | 2·3 |
| Delivery Cost (Year>1) | 130 | 1·8 |
| Communication Cost (Year 1) | 52 | 0·7 |
| Communication Cost (Year>1) | 11 | 0·2 |
| Single-dose cost (Year 1) | 556 | 7·5 |
| Single-dose cost (Year >1) | 475 | 6·4 |

**Table A.12: Extrapolation delivery costs of Sikkim to India based on costs of childhood vaccination programme.**

| State | Delivery Costs (childhood vaccination) | Population 0-6 census 2011 | Population (%) | Cost per child | Multiplier Relative difference compared to Sikkim | Delivery Cost single-dose (year 1) | Delivery Cost (year >1) |
|---|---|---|---|---|---|---|---|
| Andaman and Nicobar Islands | 79 624 | 40 878 | 0··02% | 1··95 | 0·68 | 116 | 89 |
| Andhra Pradesh | 27 245 925 | 9 142 802 | 5·56% | 2·98 | 1·05 | 178 | 136 |
| Arunachal Pradesh | 738 436 | 212 188 | 0·13% | 3·48 | 1·22 | 208 | 159 |
| Assam | 17 402 022 | 4 638 130 | 2·82% | 3·75 | 1·32 | 224 | 171 |
| Bihar | 85 989 369 | 19 133 964 | 11·63% | 4·49 | 1·58 | 269 | 205 |
| Chandigarh | 418 226 | 119 434 | 0·07% | 3·50 | 1·23 | 209 | 160 |
| Chhattisgarh | 17 061 430 | 3 661 689 | 2·23% | 4·66 | 1·64 | 279 | 212 |
| Dadra and Nagar Haveli | 229 573 | 50 895 | 0·03% | 4·51 | 1·58 | 270 | 206 |
| Daman and Diu | 106 840 | 26 934 | 0·02% | 3·97 | 1·39 | 237 | 181 |
| Goa | 772 667 | 144 611 | 0·09% | 5·34 | 1·88 | 319 | 244 |
| Gujarat | 31 427 760 | 7 777 262 | 4·73% | 4·04 | 1·42 | 242 | 184 |
| Haryana | 16 896 497 | 3 380 721 | 2·05% | 5·00 | 1·75 | 299 | 228 |
| Himachal Pradesh | 3 267 078 | 777 898 | 0·47% | 4·20 | 1·47 | 251 | 191 |
| Jammu and Kashmir | 5 961 008 | 2 018 905 | 1·23% | 2·95 | 1·04 | 176 | 135 |
| Jharkhand | 29 000 885 | 5 389 495 | 3·28% | 5·38 | 1·89 | 322 | 245 |
| Karnataka | 32 025 944 | 7 161 033 | 4·35% | 4·47 | 1·57 | 267 | 204 |
| Kerala | 11 352 691 | 3 472 955 | 2·11% | 3·27 | 1·15 | 195 | 149 |
| Lakshadweep | 36 926 | 7255 | 0·00% | 5·09 | 1·79 | 304 | 232 |
| Madhya Pradesh | 52 188 601 | 10 809 395 | 6·57% | 4·83 | 1·70 | 289 | 220 |
| Maharashtra | 65 176 224 | 13 326 517 | 8·10% | 4·89 | 1·72 | 292 | 223 |
| Manipur | 1 450 057 | 375 357 | 0·23% | 3·86 | 1·36 | 231 | 176 |
| Meghalaya | 2 083 870 | 568 536 | 0·35% | 3·67 | 1·29 | 219 | 167 |
| Mizoram | 630 416 | 168 531 | 0·10% | 3·74 | 1·31 | 224 | 171 |
| Nagaland | 761 581 | 291 071 | 0·18% | 2·62 | 0·92 | 156 | 119 |
| Delhi | 7 757 927 | 2 012 454 | 1·22% | 3·85 | 1·35 | 230 | 176 |
| Odisha | 20 326 029 | 5 273 194 | 3·21% | 3·85 | 1·35 | 230 | 176 |
| Puducherry | 776 065 | 132 858 | 0·08% | 5·84 | 2·05 | 349 | 266 |
| Punjab | 12 476 012 | 3 076 219 | 1·87% | 4·06 | 1·42 | 242 | 185 |
| Rajasthan | 43 844 044 | 10 649 504 | 6·47% | 4·12 | 1·45 | 246 | 188 |
| *Sikkim* | *182 609* | *64 111* | *0·04%* | *2·85* | *1·00* | *170* | *130* |
| Tamil Nadu | 52 091 532 | 7 423 832 | 4·51% | 7·02 | 2·46 | 419 | 320 |
| Tripura | 1 170 936 | 458 014 | 0·28% | 2·56 | 0·90 | 153 | 117 |
| Uttar Pradesh | 140 126 528 | 30 791 331 | 18·72% | 4·55 | 1·60 | 272 | 207 |
| Uttarakhand | 6 367 132 | 1355 814 | 0·82% | 4·70 | 1·65 | 281 | 214 |
| West Bengal | 50 203 695 | 10 581 466 | 6·43% | 4·74 | 1·67 | 284 | 216 |
| *India* | *737 626 159* | *164 515 253* | *100·00%* | *4·48* | *1·57* | *268* | *204* |

#### A.4. Vaccination Scenarios

The vaccination scenarios were based on data from the IARC India trial [40] and are extensively described in Man et al.[2] The scenarios are shown in Tables A.13 and A.14.

**Table A.13: Overview of single-dose waning scenario parameters.** [a]

| Assumption | HPV 16 | | | HPV 18 | | | HPV 31/33/45 | | |
|---|---|---|---|---|---|---|---|---|---|
| | $VE_{initial}$ | $VE_{plateau}$ | $rate$ | $VE_{initial}$ | $VE_{plateau}$ | $rate$ | $VE_{initial}$ | $VE_{plateau}$ | $rate$ |
| A | 0.95 | 0.95 | 0 | 0.95 | 0·95 | 0 | 0.09 | 0.09 | 0 |
| B | 0.95 | 0.60 | 0.02 | 0.95 | 0·45 | 0.02 | 0.09 | 0.45/0.95*0.09 | 0.02 |
| C | 0.90 | 0.55 | 0.02 | 0.85 | 0·35 | 0.04 | 0.09 | 0.35/0.85*0.09 | 0.04 |
| D | 0.85 | 0.50 | 0.02 | 0.55 | 0·25 | 0.08 | 0.09 | 0.25/0.55*0·09 | 0.08 |

[a] The assumptions of single-dose initial protection were derived based on the lower bound of the vaccine efficacy estimate of the IARC India vaccine trial. Waning of single-dose vaccine protection was informed by trial immunogenicity data based on the time until the antibodies levels of HPV 16/18 have decreased below different antibody detection thresholds. [9,16] The following parametric form was used for the decrease of vaccine efficacy: $(VE_{initial} - VE_{plateau}) * e^{-rate*time} + VE_{plateau}$, with $time$ in years. Vaccine protection per HPV strain are shown for a) period immediately after vaccination ($VE_{initial}$) ; and b) in the long term, ie, approximately 50 years after vaccination ($VE_{plateau}$).

**Table A.14: Vaccination Scenarios.**

| Name | Single-dose protection duration assumption* | Routine coverage | Number of scenarios |
|---|---|---|---|
| A90 (base-case) | Life-long (A) | 90% | 1 |
| A60, A70, A80, A100 | Life-long (A) | 60, 70, 80, 100% | 4 |
| B60, B70, B80, B90, B100 | Weak waning (B) | 60, 70, 80, 90, 100% | 5 |
| C60, C70, C80, C90, C100 | Intermediate waning (C) | 60, 70, 80, 90, 100% | 5 |
| D60, D70, D80, D90, D100 | Worst-case waning (D) | 60, 70, 80, 90, 100% | 5 |
| A90.CU15, A90.CU15 | Life-long (A) | 90% (CU: 60%, 90%) | 2 |
| A90.CU20, A90.CU20 | Life-long (A) | 90% (CU: 60%, 90%) | 2 |
| D90.CU20, D90.CU20 | Worst-case waning (D) | 90% (CU: 60%, 90%) | 2 |

#### A.5 Computation of Model Outcomes

##### A.5.1 Cancer Incidence

In section A.1.3 we described how the model was calibrated to national cancer incidence patterns. In projecting outcomes of vaccination, we ran simulations with two submodels, one representative of the low cancer incidence pattern (West Bengal) and one representative of the high cancer incidence pattern (Tamil Nadu). We extrapolated cancer incidence to all other states, using the results of the clustering exercise [9] shown in Table A.15. The main idea is to compute the ratio of cancer incidence per age group (denoted with index $a$) between the Indian state of interest ($cancerobs_{s,a}$) and the representative state of the cluster (either Tamil Nadu or West Bengal, $cancerobs_{cluster,a}$). Then we multiply this ratio by the predicted cancer incidence ($cancerpred_{cluster,a,y}$). For this we used the following procedure:

For each state $s$ do:

Step 1: Verify to which cluster state $s$ belongs : cluster = [High, Low]

Step 2: Select the cancer model projection ($cancerpred_{cluster}$) corresponding to $cluster$

For each age group $a$ do:

Step 3: $cancerinc_{s,a,y} = \left(\frac{cancerobs_{s,a}}{cancerobs_{\text{cluster,a}}}\right) cancerpred_{cluster,a,y}$ .

End loop $a$

End loop $s$

To obtain the Indian cancer incidence we do the following step,

Step 4: $indiancancerinc_{a,y} = \sum_s cancerinc_{s,a,y}\, weightpop_s$ ,

Where $weightpop_s$ denotes the weight of state $s$ in the total Indian population. We also applied an age adjustment in order to obtain cancer incidence by year ($inc_y$), which is necessary to compute the ICER,

Step 5: $inc_y = \sum_a indiancancerinc_{a,y}\, weightage_a$

**Table A.15: Age-specific cervical cancer incidence (per 100,000 women) by Indian state.** [a]

| State/group of states * | Source | Cluster | Age group 15-19 | 20-24 | 25-29 | 30-34 | 35-39 | 40-44 | 45-49 | 50-54 | 55-59 | 60-64 | 65-69 | 70-74 | 75-79 | 80-84 |
|---|---|---|---|---|---|---|---|---|---|---|---|---|---|---|---|---|
| Andhra Pradesh | Extracted | Low | 0 | 0·1 | 1·5 | 3·6 | 11·2 | 15·7 | 20·3 | 35·3 | 44·6 | 51 | 43·8 | 55·7 | 25·7 | 12·8 |
| Assam | Extracted | Low | 0 | 0·2 | 1·5 | 5·1 | 7·9 | 15·1 | 24·1 | 27·9 | 28·2 | 33·2 | 36·1 | 25·5 | 13·9 | 7 |
| Bihar | Inferred | Low | 0 | 0·3 | 1 | 4·3 | 8·4 | 16·2 | 22·2 | 28·6 | 30·7 | 34·9 | 31·9 | 31·9 | 15·3 | 7·6 |
| Chhattisgarh | Inferred | Low | 0 | 0·3 | 1 | 4·3 | 8·4 | 16·2 | 22·2 | 28·6 | 30·7 | 34·9 | 31·9 | 31·9 | 15·3 | 7·6 |
| Delhi | Extracted | High | 0 | 0·6 | 1·2 | 4·4 | 11·2 | 21·8 | 32·3 | 38·5 | 45·4 | 62·3 | 57 | 51·4 | 31·9 | 16 |
| Goa + Daman & Diu | Inferred | Low | 0 | 0·3 | 1 | 4·3 | 8·4 | 16·2 | 22·2 | 28·6 | 30·7 | 34·9 | 31·9 | 31·9 | 15·3 | 7·6 |
| Gujarat + Dadra & Nagar Haveli | Extracted | Low | 0 | 0 | 0·8 | 7·9 | 14 | 24·2 | 19·7 | 34·4 | 19·6 | 30·6 | 25·7 | 30·6 | 13·6 | 6·8 |
| Haryana | Inferred | Low | 0 | 0·3 | 1 | 4·3 | 8·4 | 16·2 | 22·2 | 28·6 | 30·7 | 34·9 | 31·9 | 31·9 | 15·3 | 7·6 |
| Himachal Pradesh | Inferred | Low | 0 | 0·3 | 1 | 4·3 | 8·4 | 16·2 | 22·2 | 28·6 | 30·7 | 34·9 | 31·9 | 31·9 | 15·3 | 7·6 |
| Jammu & Kashmir | Inferred | Low | 0 | 0·3 | 1 | 4·3 | 8·4 | 16·2 | 22·2 | 28·6 | 30·7 | 34·9 | 31·9 | 31·9 | 15·3 | 7·6 |
| Jharkhand | Inferred | Low | 0 | 0·3 | 1 | 4·3 | 8·4 | 16·2 | 22·2 | 28·6 | 30·7 | 34·9 | 31·9 | 31·9 | 15·3 | 7·6 |
| Karnataka | Extracted | High | 0 | 0·2 | 0·9 | 4·9 | 10 | 23·8 | 40·4 | 52·2 | 63·7 | 64 | 79·6 | 74·1 | 30·2 | 15·1 |
| Kerala + Lakshadweep | Extracted | Low | 0 | 0·2 | 0·4 | 0·3 | 2·5 | 9·2 | 15 | 19·1 | 30 | 38·4 | 31·9 | 39·3 | 11·7 | 5·8 |
| Madhya Pradesh | Extracted | High | 0·2 | 0·2 | 2 | 4·3 | 12·8 | 23·8 | 30·4 | 53·3 | 48·8 | 65 | 61·8 | 76·4 | 17·7 | 8·8 |
| Maharashtra | Extracted | Low | 0 | 0·4 | 1·3 | 3·8 | 10·8 | 20·8 | 27·1 | 35·4 | 36·8 | 50·5 | 51·7 | 43·3 | 22·4 | 11·2 |
| Manipur | Extracted | Low | 0 | 0·6 | 0·1 | 4·2 | 7·4 | 12·2 | 14·8 | 21·3 | 25·1 | 35 | 32·7 | 39·4 | 23·2 | 11·6 |
| Orissa | Inferred | Low | 0 | 0·3 | 1 | 4·3 | 8·4 | 16·2 | 22·2 | 28·6 | 30·7 | 34·9 | 31·9 | 31·9 | 15·3 | 7·6 |
| Other North Eastern States § | Extracted | High | 0 | 1·1 | 1·9 | 10·7 | 20·3 | 40·9 | 50 | 54·3 | 52·2 | 47·8 | 31·9 | 44 | 16·6 | 8·3 |
| Punjab + Chandigarh | Extracted | High | 0 | 0·1 | 2·2 | 3 | 13 | 23·8 | 32·3 | 47·2 | 47·8 | 48·7 | 52·8 | 35·2 | 33·3 | 16·6 |
| Rajasthan | Inferred | Low | 0 | 0·3 | 1 | 4·3 | 8·4 | 16·2 | 22·2 | 28·6 | 30·7 | 34·9 | 31·9 | 31·9 | 15·3 | 7·6 |
| Sikkim | Extracted | Low | 0 | 0 | 0·7 | 4·3 | 12 | 26·1 | 29 | 24·6 | 42·8 | 26·5 | 20·8 | 12·4 | 0 | 0 |
| Tamil Nadu + Puducherry | Extracted | High | 0 | 0·4 | 0·9 | 4·3 | 13·4 | 33·7 | 48·1 | 60·8 | 68·9 | 75·5 | 67·1 | 65·1 | 19·9 | 9·9 |
| Uttar Pradesh | Inferred | Low | 0 | 0·3 | 1 | 4·3 | 8·4 | 16·2 | 22·2 | 28·6 | 30·7 | 34·9 | 31·9 | 31·9 | 15·3 | 7·6 |
| Uttarakhand | Inferred | Low | 0 | 0·3 | 1 | 4·3 | 8·4 | 16·2 | 22·2 | 28·6 | 30·7 | 34·9 | 31·9 | 31·9 | 15·3 | 7·6 |
| West Bengal + Andaman & Nicobar Islands | Extracted | Low | 0 | 0·3 | 0·6 | 3·5 | 6·2 | 15·9 | 23·7 | 27·5 | 26·7 | 29·1 | 34·8 | 31·7 | 21·5 | 10·8 |

[a] Other North Eastern States include Arunachal Pradesh, Nagaland, Meghalaya, Mizoram, and Tripura.

[b] Extracted from CI5 or NCDIR or inferred based on clustering framework (“footprinting”) described in Man et al 2022.[9]

##### A.5.2 Disability adjusted life years

Disability weights and their durations are shown in Table A.15 and are based on Global Burden and Disease (GBD) 2017 update, adapted from Abbas et al.[41] Life years are calculated as the total number of persons alive in a year ($LY_{a,y}$). In order to obtain disability adjusted life years per year ($y$) and age group ($a$) ($DALY_{a,y}$) we subtract the unweighted life years lived with disability ($YLDunw$), and add weighted years lived with disability, with weights and durations given in Table A.16 ($YLDw$):

$$YLDunw_{a,y} = incidence_{a,y} \;\; LY_{a,y} \; + \; prevalence_{a,y} \;\; LY_{a,y} + mortality_{a,y} \;\; LY_{a,y} \,,$$

$$YLDw_{a,y} = incidence_{a,y}\, LY_{a,y} \left( weight_{inc}\, duration_{inc} + weight_{prev} \;\left(1 - duration_{inc}\right)\right) + prevalence_{a,y} \;\; LY_{a,y}\, weight_{prev} \; + \; mortality_{a,y} \;\; LY_{a,y} \left( weight_{term}\, duration_{term} + weight_{metast}\, duration_{metast} + weight_{prev}\left(1 - \; duration_{term} - duration_{metast}\right)\right) \,,$$

$$DALY_{a,y} = LY_{a,y} \; - YLDunw_{a,y} \; + YLDw_{a,y} \,.$$

In order to calculate incremental health gains (*IH*) we sum $DALY_{a,y}$ over all age groups first,

$$tDALY_{y} = \sum_{a} DALY_{a,y} \;\; weightage_{a}.$$

Then we obtain incremental health gains as the difference in DALYs between the no vaccination scenario and the vaccination scenario of interest:

$$IH_{y,scen} = tDALY_{y,novacc} - tDALY_{y,scen}.$$

**Table A.16: Disability weights and durations for different phases of cervical cancer in GBD 2017 study. [a]**

| Health State | Weight Name | Value | Duration |
|---|---|---|---|
| Diagnosis and primary therapy phase | $weight_{inc}$ | 0·288 | 4·8 months |
| Controlled phase | $weight_{prev}$ | 0·049 | Remainder of time |
| Metastatic phase | $weight_{metast}$ | 0·451 | 9·21 months |
| Terminal phase | $weight_{term}$ | 0·54 | 1 month |

[a] Based on Abbas et al 2020.[41] GBD = Global Burden of Disease.

##### A.5.3 Costs

Costs include two main categories, costs of vaccination and costs of cervical cancer treatment. Costs of vaccination per year ($Vac_y$) are calculated by multiplying the sum of vaccine dose cost and delivery cost by the weight of girls aged 10 in the population. For two-dose we applied the two-dose cost described in Table A.11. For catch-up we use the weight in the population of girls in the catch-up cohort of interest (either 11−15 or 11−20 years). For the first two years we assume that the government buys a number of vaccine doses sufficient for the whole population, independently of the coverage, since the government is unlikely to know in advance what will be the uptake of vaccination. Yearly costs of cervical cancer treatment ($CCC_y$) are obtained by multiplying the cancer incidence (by stage) by the costs described in Table A.8. Incremental costs per year and scenario ($IC_{y,scen}$) are given by:

$$IC_{y,scen} = Vac_{y,scen} - (CCC_{y,novacc} - CCC_{y,scen}).$$

##### A.5.4 ICER

ICER (Incremental Cost-effectiveness ratio) is computed as the ratio between incremental costs and DALYs averted compared to the scenario without vaccination. These are summed over all the years of the 100-year time horizon:

$$ICER_{scen} = \frac{(\sum_y^{100} IC_{y,scen})}{(\sum_y^{100} IH_{y,scen})}.$$

#### A.6 HPV-FRAME Checklist

**Table A.17: HPV-FRAME Checklist, Core Reporting Standard.**

| **Core Reporting Standard** | | | |
|---|---|---|---|
| **Inputs** | **Reported by age?** | **Report by sex?** | **Comments** |
| Target population for intervention | Y | Y | Only vaccination in girls and women were considered. Age of routine and catch-up vaccination were reported. |
| Sexual behavior | Y | Y | HPV incidence computed based on EpiMetHeos model (See Man et al [2]) |
| Cohort examined for evaluation / time horizon | Y | Y | Multicohort model representing age distribution of Indian population; time horizon 100 years. |
| Quality of life assumptions | N | NA | Based on Global Burden of Disease 2017 [41] (Table A.15); No disability adjusted weights by age were available. |
| Calibration | Y | NA | For calibration of transmission model see Man et al.[2] Cervical Cancer progression model calibration is described in sections A1.2 and A1.3. |
| Validation (where possible) | Y | NA | Cervical cancer incidence predicted by the model checked against observed cervical cancer incidence in two Indian states, Tamil Nadu and West Bengal (Figure A.2) |
| Costs | N | NA | See Section A.3. All costs valued in USD, and discounted at 3% according to WHO guidelines. Costs also valued in IUSD as a sensitivity analysis. |

[a] Y= Yes, N=No, NA=Not Applicable.

**Table A.18: HPV-FRAME Checklist, Reporting standard for HPV vaccination in adolescent individuals.[a]**

| **Reporting standard for HPV vaccination in adolescent individuals** | | | | |
|---|---|---|---|---|
| **Inputs** | **Reported** | **Reported by age?** | **Report by sex?** | **Comments** |
| Vaccine uptake | Y | Y | Y | Single- and two-dose schedules were considered. Only vaccination in girls and women were considered. Uptake between 60-100% were considered for routine vaccination. Uptake of 60% and 90% were considered for catch-up vaccination |
| Vaccine efficacy | Y | NA | NA | Efficacy by dose schedule and HPV type was considered. Efficacy was independent of age. |
| Vaccine cross-protection | Y | Y | NA | Level of cross-protection for HPV 31/33/35 was reported. |
| Duration vaccine protection and waning | Y | N | NA | Waning assumption by dose schedule and HPV type was considered. |
| Vaccine and delivery costs | Y | NA | NA | Vaccine price is equal to GAVI price. Delivery costs extrapolated from the state of Sikkim, based on state-specific delivery costs estimates of childhood vaccination. [38] |
| Pre-vaccination disease burden (including population attributable fractions for HPV) | Y | Y | NA | Current cervical cancer incidence considered as baseline. |
| Duration of natural immunity | Y | N | NA | Natural immunity was independent of age. |
| **Outputs** | **Reported** | **Reported by age?** | **Report by sex?** | **Report as calibration or validation target? (Y/N)** |
| Absolute reductions in HPV infections, and/or warts, post-vaccination | N | N | N | Reduction in HPV infection reported in Man et al.[2] |
| Absolute reductions in CIN2+ post-vaccination | N | N | N | CIN2+ reduction is not reported as it is not detected in a context without screening. |
| Absolute reductions in invasive cancer (cervical and other HPV cancers, as relevant) | Y, for cervical cancer | N | NA | Yes, it was a validation target for cancer progression model calibration. (See Figure A.2) |

[a] Y= Yes, N=No, NA=Not Applicable, F= Female, ICER= Incremental Cost Effectiveness Ratio.

**Table A.19: HPV-FRAME Checklist, Reporting standard for models of HPV prevention in LMIC.**

| Inputs | Reported | Reported by age? | Report by sex? | Comments |
|---|---|---|---|---|
| HIV prevalence rates if endemic in country | N | N | NA | The effects of HIV are not modelled because of the low cervical cancer attributable fraction due to HIV in the whole of India. [42] |
| Description of any opportunistic or pilot/demonstration screening project ongoing | Y | N | NA | We assume no screening in this study, which is justified by the very low screening coverage (2% in women aged 35−49 years) in India |

**Table A.20: HPV-FRAME Checklist, Reporting standards for evaluations assessing alternative vaccine types or reduced-dose schedules.**

| Inputs | Reported | Reported by age? | Report by sex? | Comments |
|---|---|---|---|---|
| Vaccine efficacy/waning | Y | N | NA | See Table A.17 |
| Timing between doses (for 2-dose) | NA | NA | NA | Timing between the two doses under two-dose vaccination schedule was not modelled. |
| Vaccine cross-protection | Y | N | NA | See Table A.17 |
| Cost | Y | NA | NA | Costs per single-dose and two-dose schedule are reported in Table A.11. Willlingness to pay threshold based on WHO guideline (100% GDP per capita) and on Jit 2021 [43] (30% GDP per capita); |
| **Outputs** | **Reported** | **Reported by age?** | **Report by sex?** | **Report as calibration or validation target (Y/N)?** |
| Threshold cost per dose | N | N | N | |

#### A.7 Details on Sensitivity Analyses

##### A.7.1 Vaccine cost reduction over time

The idea of this sensitivity analysis is to compute the ICER under a scenario where vaccine cost reduces over time, where the rate of price reduction is informed by past price reductions for GAVI-supported pneumococcal vaccine.[44] The rate of vaccine price reduction was calculated based on the observed price reduction between 2010 and 2019. In 2010 ("First Supply Agreement") the contracted vaccine price per was $3·5 USD.This reduced incrementally to $2·9 USD in 2019 ("Fifth Supply Agreement"). The rate of price reduction per year equals 2%. For this sensitivity analysis we assume that this rate of vaccine price reduction would also occur for HPV vaccine.

##### A.7.2 Distributions of parameter values for probabilistic analysis

For this analysis we only included a few influential model parameters. It would be unfeasible to include all model parameters, as this would make this task computationally prohibitive given the high number of simulation runs per scenario required. Where available we used published 95% confidence intervals for the model parameters included in the analysis, namely for the durations from CIN2/3 to cervical cancer. For other parameters, namely, for costs, we used the heterogeneity in the data to construct a 95% confidence interval for each model parameter. The parameters for the sampling distributions of each parameter are obtained via simulation based on the lower and higher bounds of the confidence interval (shown in Table A.20).

**Table A.21: Sampling distributions for model parameters included in probabilistic analysis.**

| Model Parameter | Values for mean and uncertainty bounds | Sampling Distribution | Source |
|---|---|---|---|
| Duration to Cancer | | | |
| $k_{16}, k_{18}, k_{other}$ | Mean: 9·67 ; 9·67 ; 2·49<br>Lower bound: 4·74 ; 4·74 ; 1·54<br>Higher bound: 24·0 ; 24·0, 3·95 | *Gamma* | Confidence intervals in Vink et al 2013 [8] |
| $\theta_{16}, \theta_{18}, \theta_{other}$ | Mean: 3·33 ; 3·33 ; 9·14<br>Lower bound: 2·28 ; 2·28 ; 5·91<br>Higher Bound: 5·06 ; 5·06 ; 13·6 | *Gamma* | Confidence intervals in Vink et al 2013 [8] |
| Survival | | | |
| 5-year Cancer Survival | Mean: 0·55<br>Lower Bound: 0·35<br>Higher Bound: 0·67 | *Beta* | See Table A.4. Confidence interval based on lower/highest survival. |
| Costs | | | |
| Radical Hysterectomy | Mean: 80 392<br>Lower Bound: 64 994<br>Higher Bound: 125 000 | *Lognormal* | Lower and upper bounds of the confidence interval based on hospitals with low/highest cost (Table A.6) |
| Chemo-radiotherapy | Mean: 72 233<br>Lower Bound: 40 000<br>Higher Bound: 90 405 | *Lognormal* | Lower and upper bounds of the confidence interval based on hospitals with low/highest cost (Table A.6) |
| Delivery Costs (Year 1, Year > 1) | Mean: 320, 216<br>Lower Bound: 230, 147<br>Higher Bound: 374, 256 | *Lognormal* | Lower and upper bounds of the confidence interval based on states with 2nd highest and 2nd lowest delivery cost (i.e., 5th and 95th percentile).[38] |
| Cost multiplier private vs public | Mean: 4·1<br>Lower Bound: 3·6<br>Higher Bound: 5·3 | *Lognormal* | Lower and upper bounds of the confidence interval based on urban and rural private vs public cost multipliers. [32] |

**Supplementary Appendix B**

#### Additional Model Results for: Health economic impact of introducing single-dose HPV vaccination in India.

de Carvalho TM, Man I, Georges D, Saraswati LR, Bhandari P, Kataria I, Siddiqui M, Muwonge R, Lucas E, Sankaranarayanan R, Basu P, Berkhof J, Bogaards JA, Baussano I.

**List of figures**

Figure B.1: Life years gained.

Figure B.2: Projected cervical cancer incidence reduction after introduction of single-dose HPV vaccination in India for vaccine protection assumption A (no waning).

Figure B.3: Projected cervical cancer incidence reduction after introduction of single-dose HPV vaccination in India for vaccine protection assumption B.

Figure B.4: Projected cervical cancer incidence reduction after introduction of single-dose HPV vaccination in India for vaccine protection assumption C.

Figure B.5: Projected cervical cancer incidence reduction after introduction of single-dose HPV vaccination in India for vaccine protection assumption D.

Figure B.6: Projected cervical cancer incidence reduction after introduction of single-dose HPV vaccination in India by catch-up scenario.

Figure B.7: Undiscounted incremental costs and health effects of single- and two-dose vaccination versus no vaccination.

Figure B.8: Incremental Costs and incremental health effects for two-dose versus single-dose schedule.

Figure B.9: Incremental Costs and incremental health effects for catch-up scenarios under a single-dose schedule.

Figure B.10: Undiscounted incremental costs and health effects of single- and two-dose vaccination versus no vaccination ($IUSD).

Figure B.11: Incremental costs and health effects of single- and two-dose vaccination versus no vaccination ($IUSD).

Figure B.12: Incremental costs and health effects of two-dose versus single-dose vaccination (assuming lifetime protection for two-dose vaccination) ($IUSD).

Figure B.13: Health economic outcomes by catch-up scenario ($IUSD).

Figure B.14: Univariate sensitivity analysis on the ICER for cost variables ($IUSD).

**List of tables**

Table B.1 : Health economic outcomes by Indian state for the base case scenario.

#### Section 1: Life years gained and reduction in cancer incidence per scenario

**Figure B.1: Life years gained.**

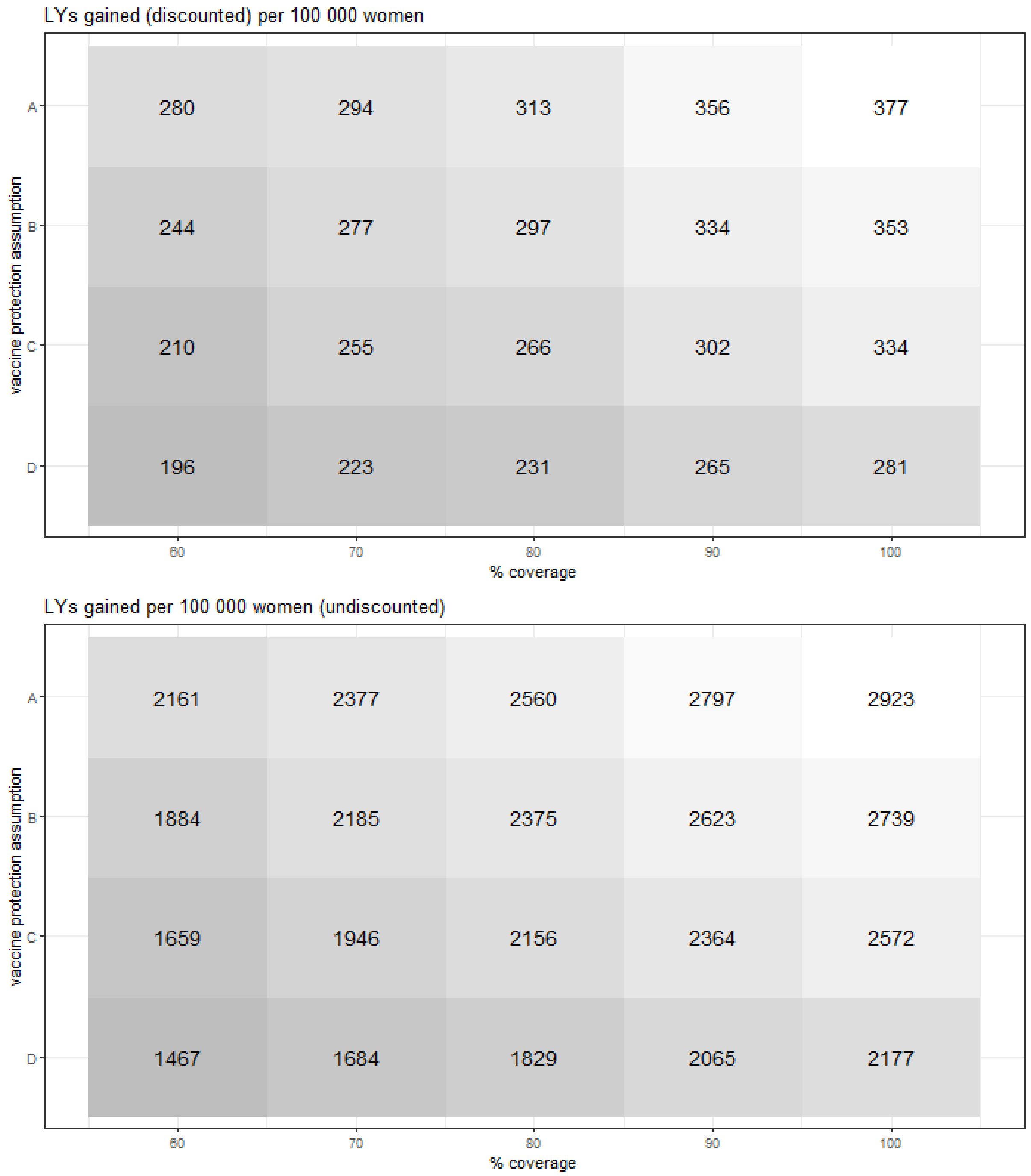

[a] Life-years gained relative to no vaccination are discounted at 3%.

**Figure B.2: Projected cervical cancer incidence reduction after introduction of single-dose HPV vaccination in India for vaccine protection assumption A (no waning).[a]**

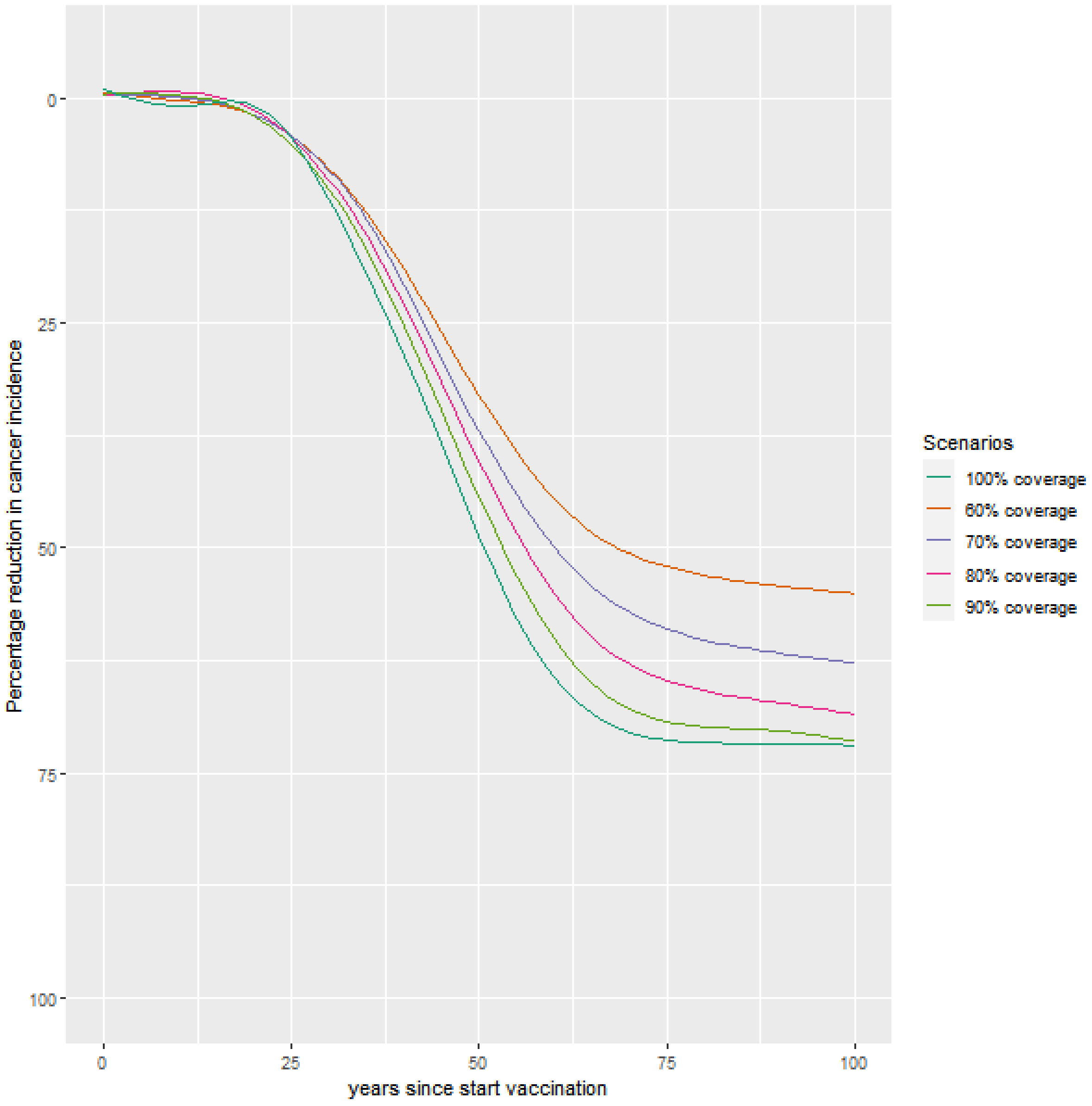

[a] Vaccine protection assumption A denotes a 95% vaccine efficacy against HPV16/18 with no waning, and 9% cross-protection for types HPV 31/33/45. Cancer incidence is age-standardised using Indian population weights.

**Figure B.3: Projected cervical cancer incidence reduction after introduction of single-dose HPV vaccination in India for vaccine protection assumption B.[a]**

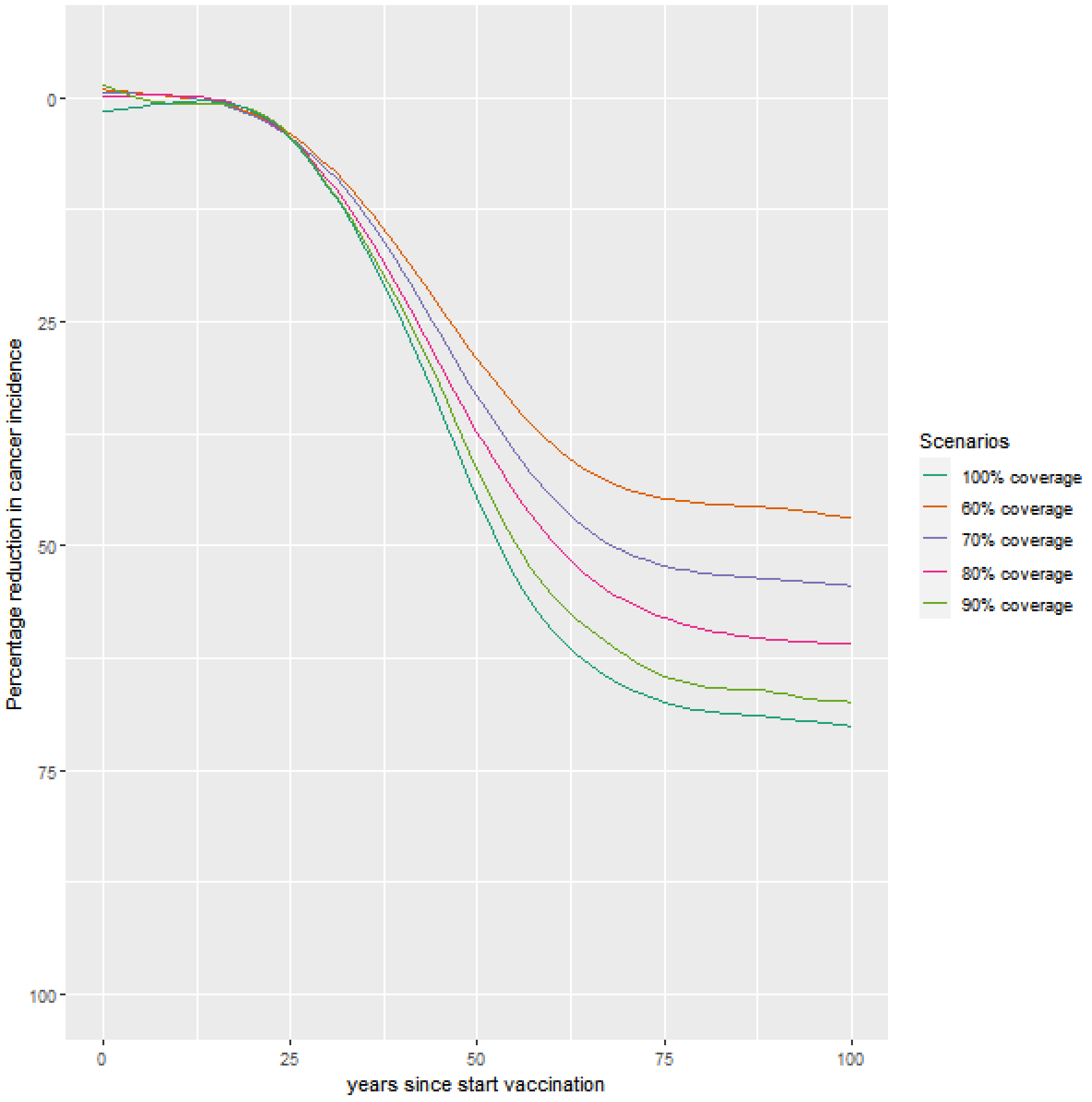

[a] Vaccine protection assumption B denotes a 95% vaccine efficacy against HPV16/18, and 9% cross-protection for types HPV 31/33/45. Vaccine protection wanes during the first 20 years since vaccination with remaining efficacy of 80% of the initial efficacy. Cancer incidence is age-standardised using Indian population weights.

**Figure B.4: Projected cervical cancer incidence reduction after introduction of single-dose HPV vaccination in India for vaccine protection assumption C.**[a]

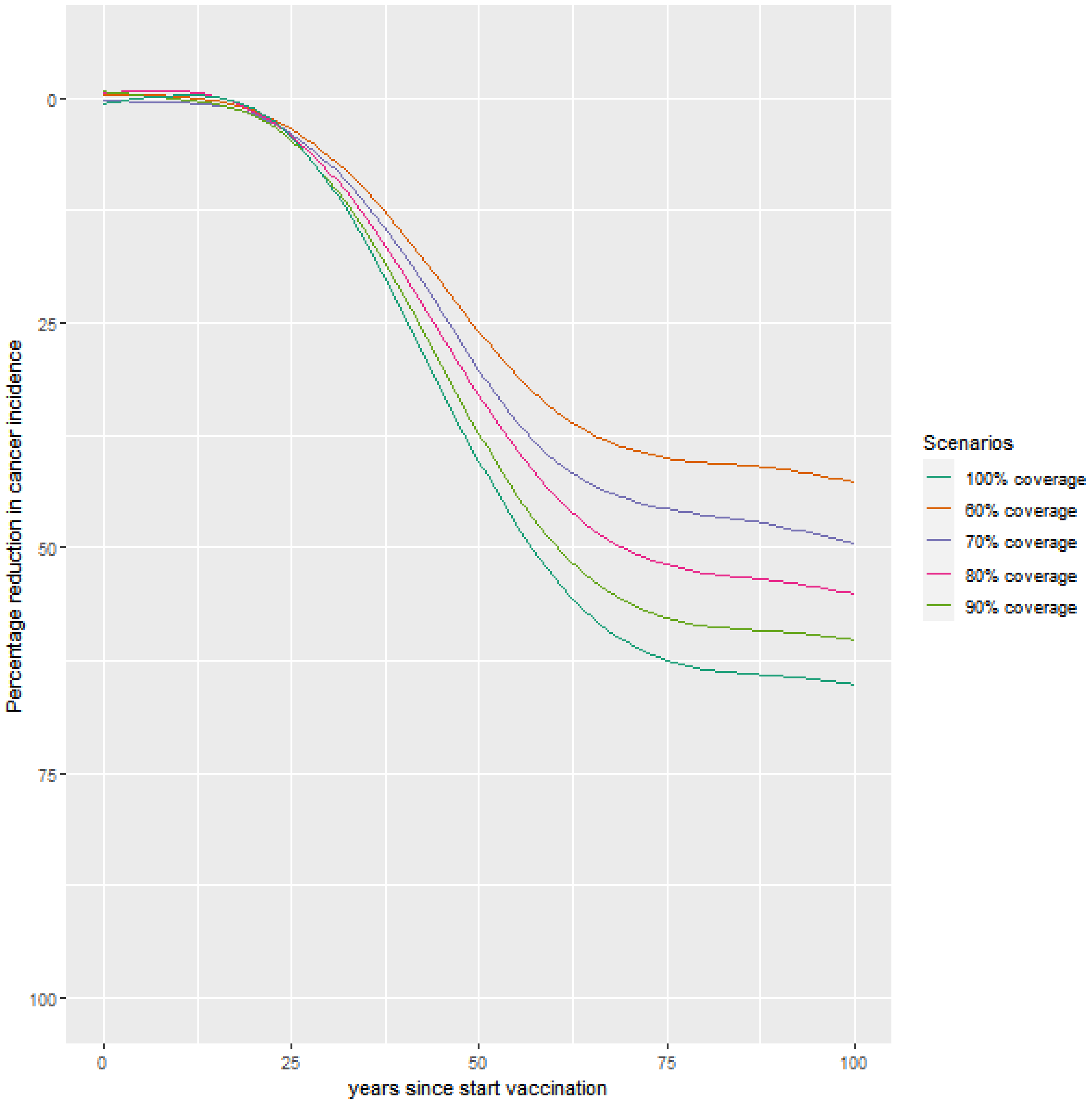

[a] Vaccine protection assumption C denotes 90% vaccine efficacy against HPV16, 85% vaccine efficacy against HPV18, with exponentially decreasing efficacy during the first 20 years since vaccination until 55% (HPV16) and 35% (HPV18). Cross-protection for types HPV 31/33/45 starts at 9% with efficacy waning at the same rate as for HPV18. Cancer incidence is age-standardised using Indian population weights.

**Figure B.5: Projected cervical cancer incidence reduction after introduction of single-dose HPV vaccination in India for vaccine protection assumption D.[a]**

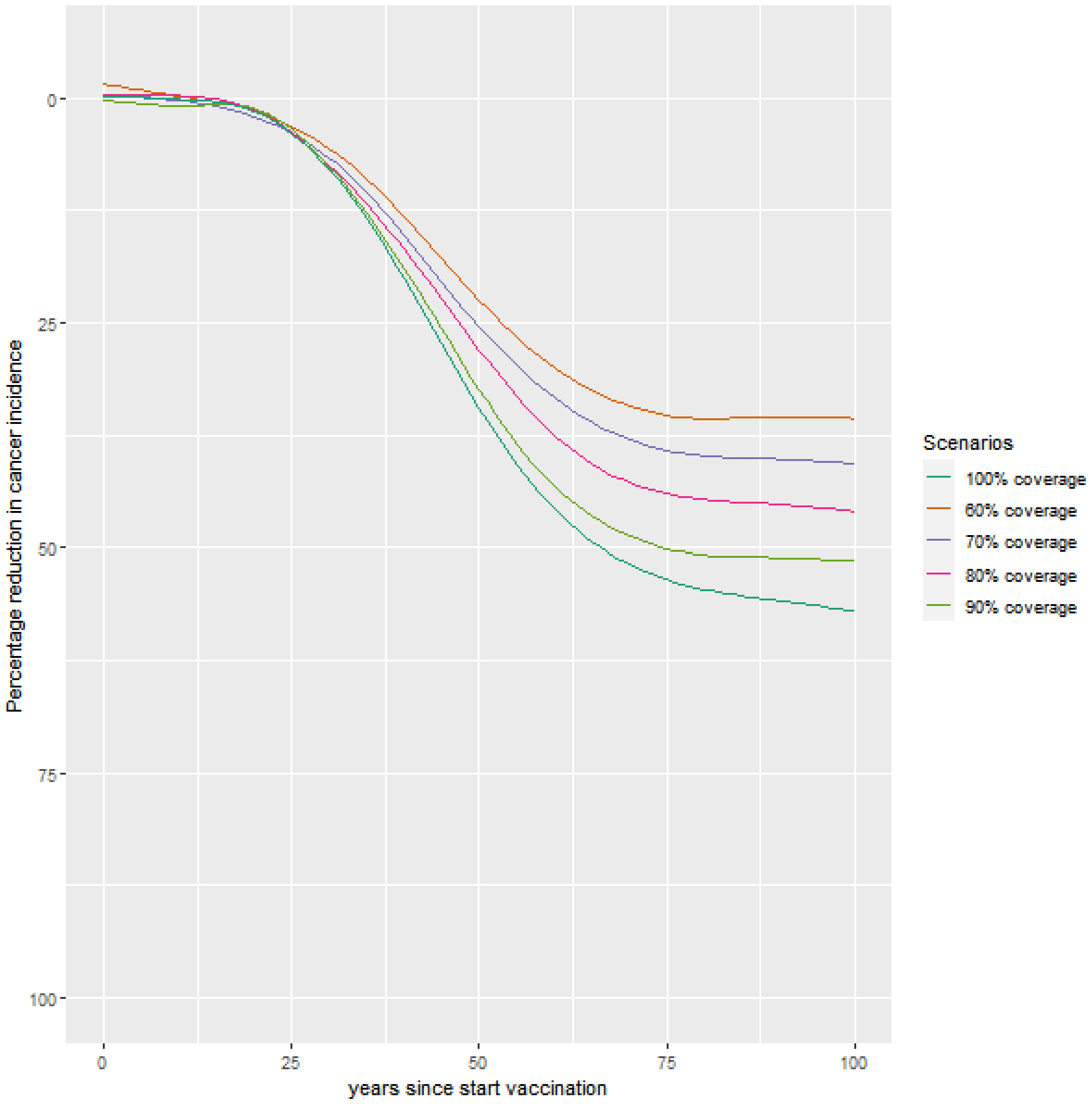

[a] Vaccine protection assumption D denotes a 85% vaccine efficacy against HPV16, 55% against HPV18 and 9% cross-protection for types HPV 31/33/45. Vaccine protection wanes during the first 20 years since vaccination with remaining efficacy of 65% of the initial efficacy. Cancer incidence is age-standardised using Indian population weights.

**Figure B.6: Projected cervical cancer incidence reduction after introduction of single-dose HPV vaccination in India by catch-up scenario.[a]**

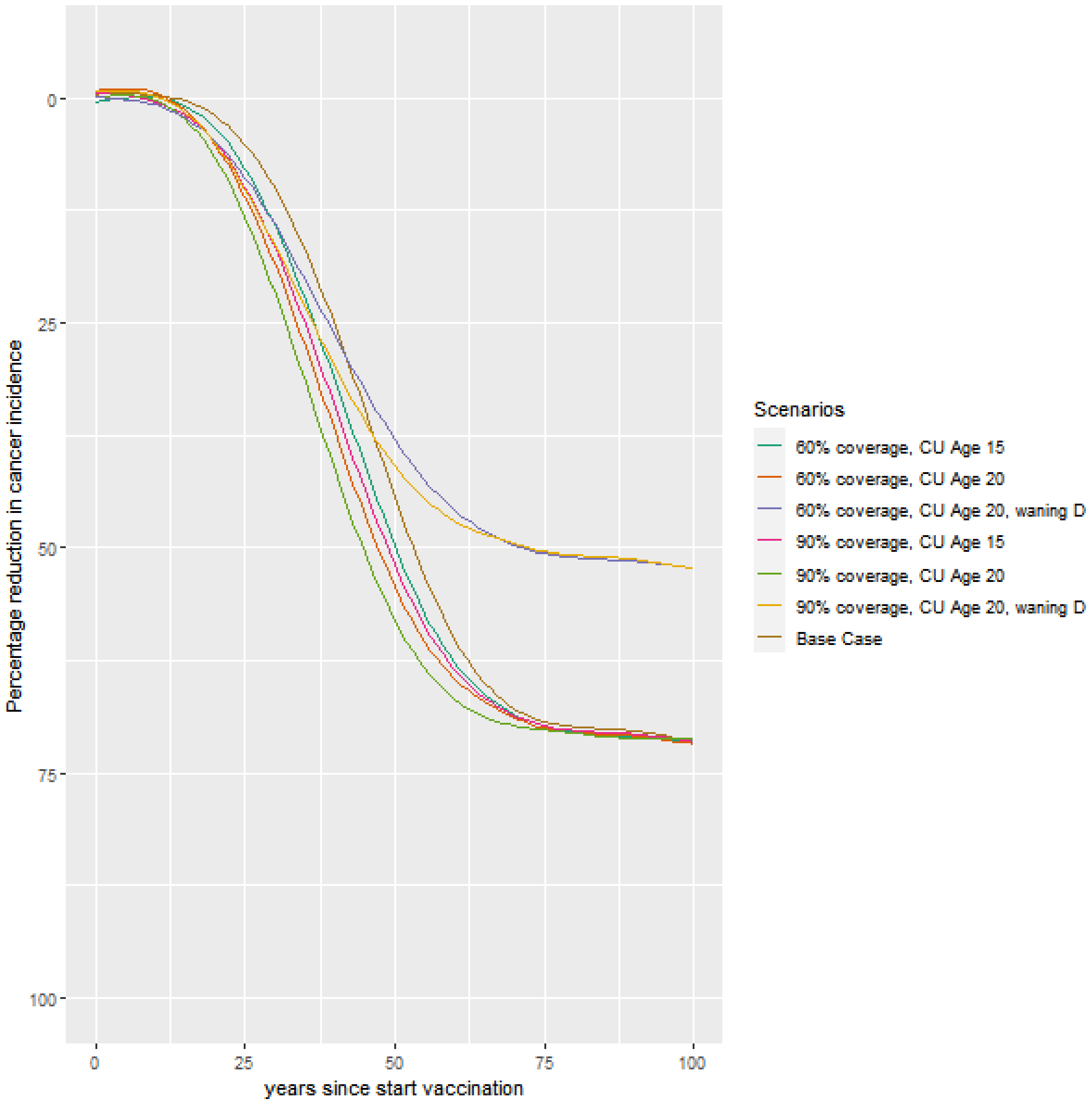

[a] Catch-up vaccination until age 15 or 20 is shown for 60% or 90% coverage catch-up coverage, 90% coverage for regular vaccinated girls and with efficacy and waning as in the Scenario A. We also show two scenarios with vaccine efficacy and waning as in Assumption D.

**Section 2: Additional Results**

**Figure B.7: Undiscounted incremental costs and health effects of single- and two-dose vaccination versus no vaccination (USD).[a]**

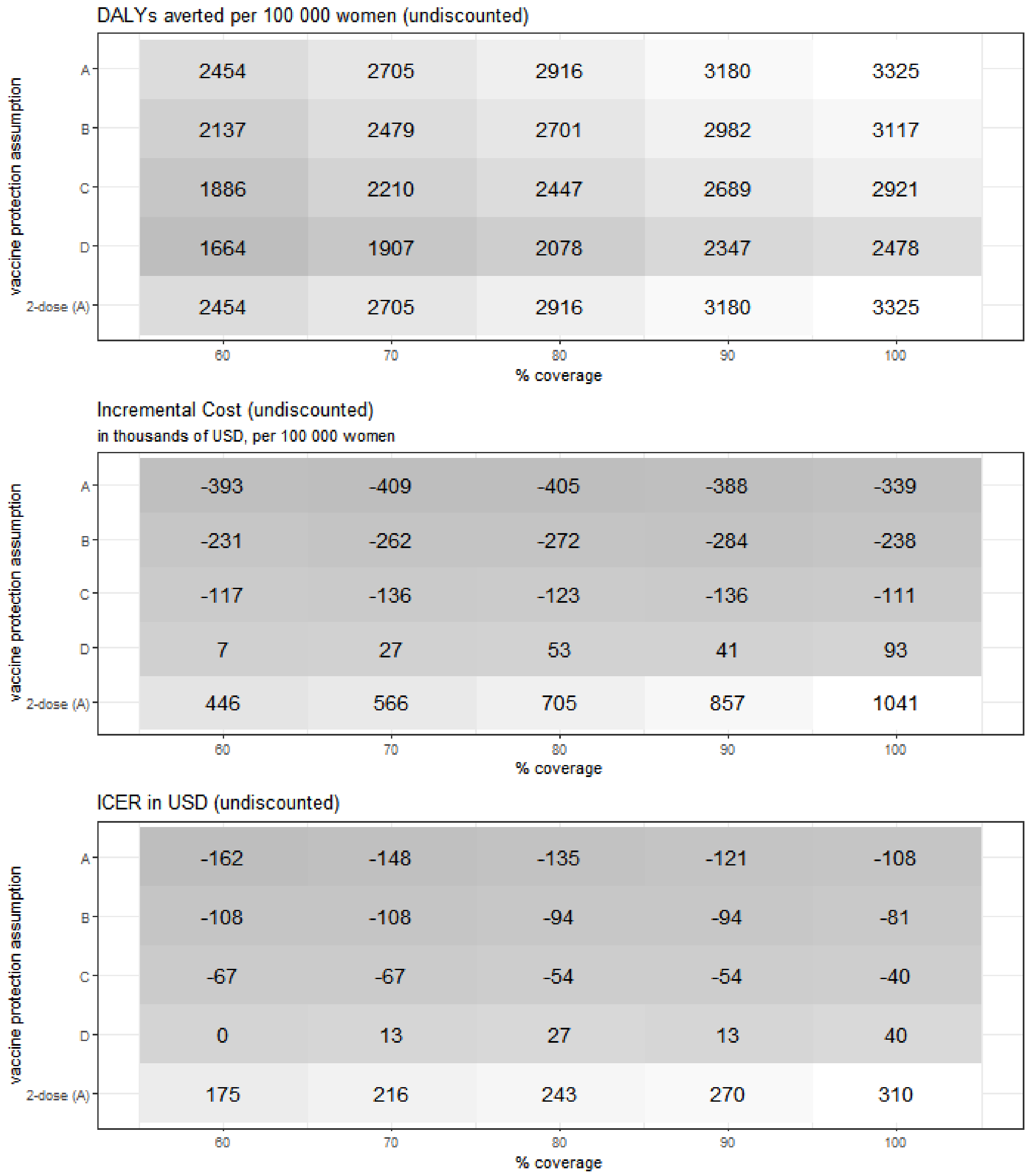

[a] Costs are given in $USD at 2020 prices. Costs and disability-adjusted life years (DALYs) averted are not discounted. ICER denotes incremental cost-effectiveness ratio per DALY averted relative to no vaccination.

**Table B.1: Health economic outcomes by Indian state for the base case scenario.** [a]

| Indian state / group of Indian states | DALYs | Incremental Costs (thousands USD) | ICER (USD) |
|---|---|---|---|
| India (all states) | 412 | 167 | 405 |
| Andhra Pradesh | 380 | 84 | 220 |
| Assam | 367 | 162 | 440 |
| Bihar | 368 | 184 | 501 |
| Chhattisgarh | 368 | 190 | 516 |
| Delhi | 549 | 158 | 288 |
| Goa + Daman & Diu | 368 | 213 | 578 |
| Gujarat + Dadra & Nagar Haveli | 374 | 142 | 378 |
| Haryana | 368 | 201 | 547 |
| Himachal Pradesh | 368 | 175 | 475 |
| Jammu & Kashmir | 368 | 134 | 363 |
| Jharkhand | 368 | 214 | 581 |
| Karnataka | 559 | 140 | 251 |
| Kerala + Lakshadweep | 353 | 207 | 587 |
| Madhya Pradesh | 555 | 169 | 305 |
| Maharashtra | 382 | 139 | 364 |
| Manipur | 359 | 200 | 556 |
| Orissa | 368 | 163 | 444 |
| Other North Eastern States | 572 | 40 | 70 |
| Punjab + Chandigarh | 551 | 158 | 287 |
| Rajasthan | 368 | 172 | 467 |
| Sikkim | 376 | 99 | 264 |
| Tamil Nadu + Puducherry | 569 | 191 | 336 |
| Uttar Pradesh | 368 | 186 | 506 |
| Uttarakhand | 368 | 191 | 519 |
| West Bengal + Andaman & Nicobar Islands | 364 | 210 | 577 |

[a] Base-case scenario denotes single-dose vaccination with 90% uptake among 10-year-old girls, a 95% vaccine efficacy against HPV16/18 with no waning, and 9% cross-protection for types HPV 31/33/45. Costs are given in $USD at 2020 prices. Costs and disability-adjusted life years (DALYs) averted are discounted at 3%. ICER denotes incremental cost-effectiveness ratio per DALY averted relative to no vaccination.

**Figure B.8: Incremental Costs and incremental health effects for two-dose versus single-dose schedule.[a]**

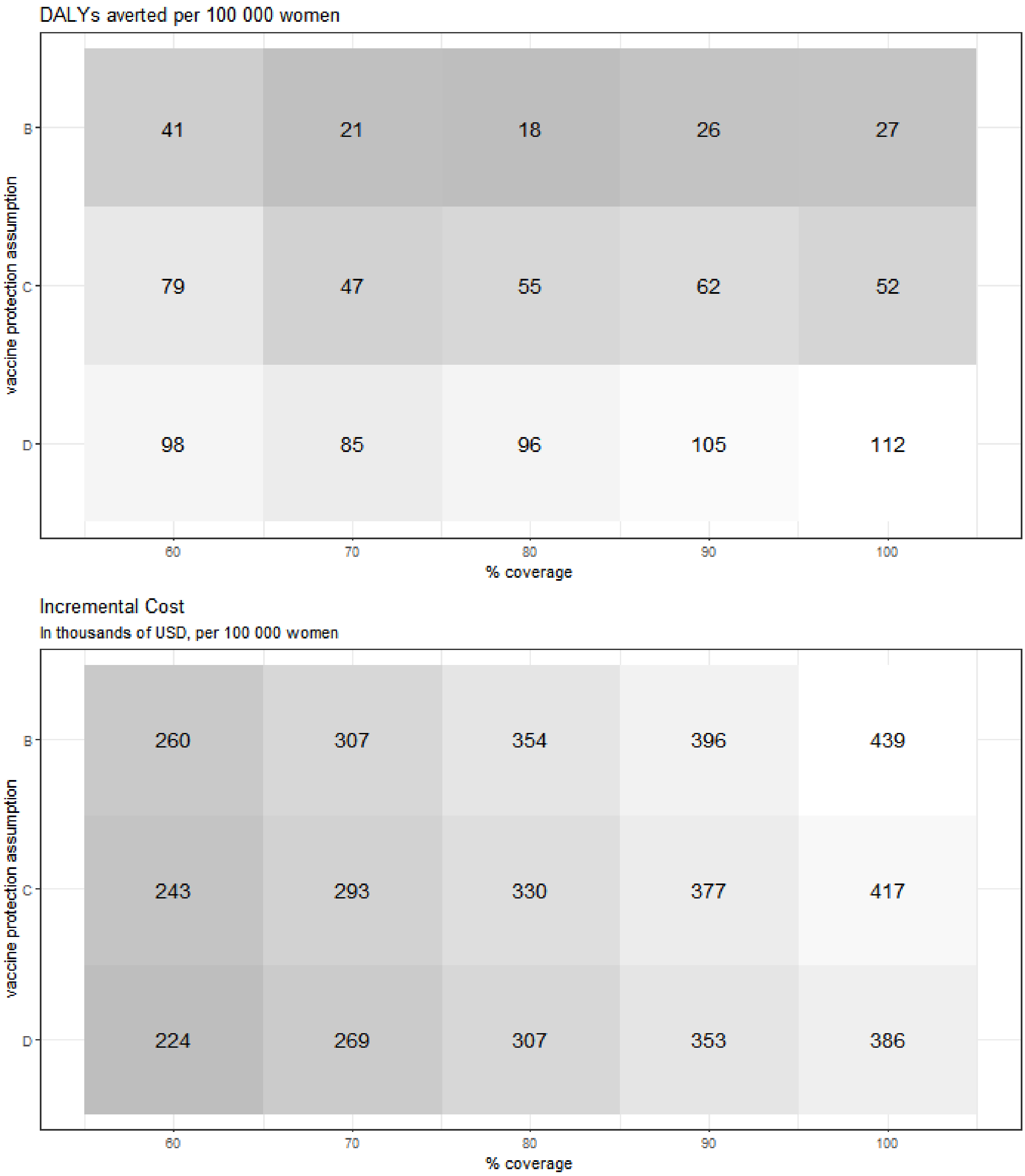

[a] Costs are given in $USD at 2020 prices. Costs and disability-adjusted life years (DALYs) averted are discounted at 3%. See Figure 3 for the corresponding ICER.

**Figure B.9: Incremental Costs and incremental health effects for catch-up scenarios under a single-dose schedule.[a]**

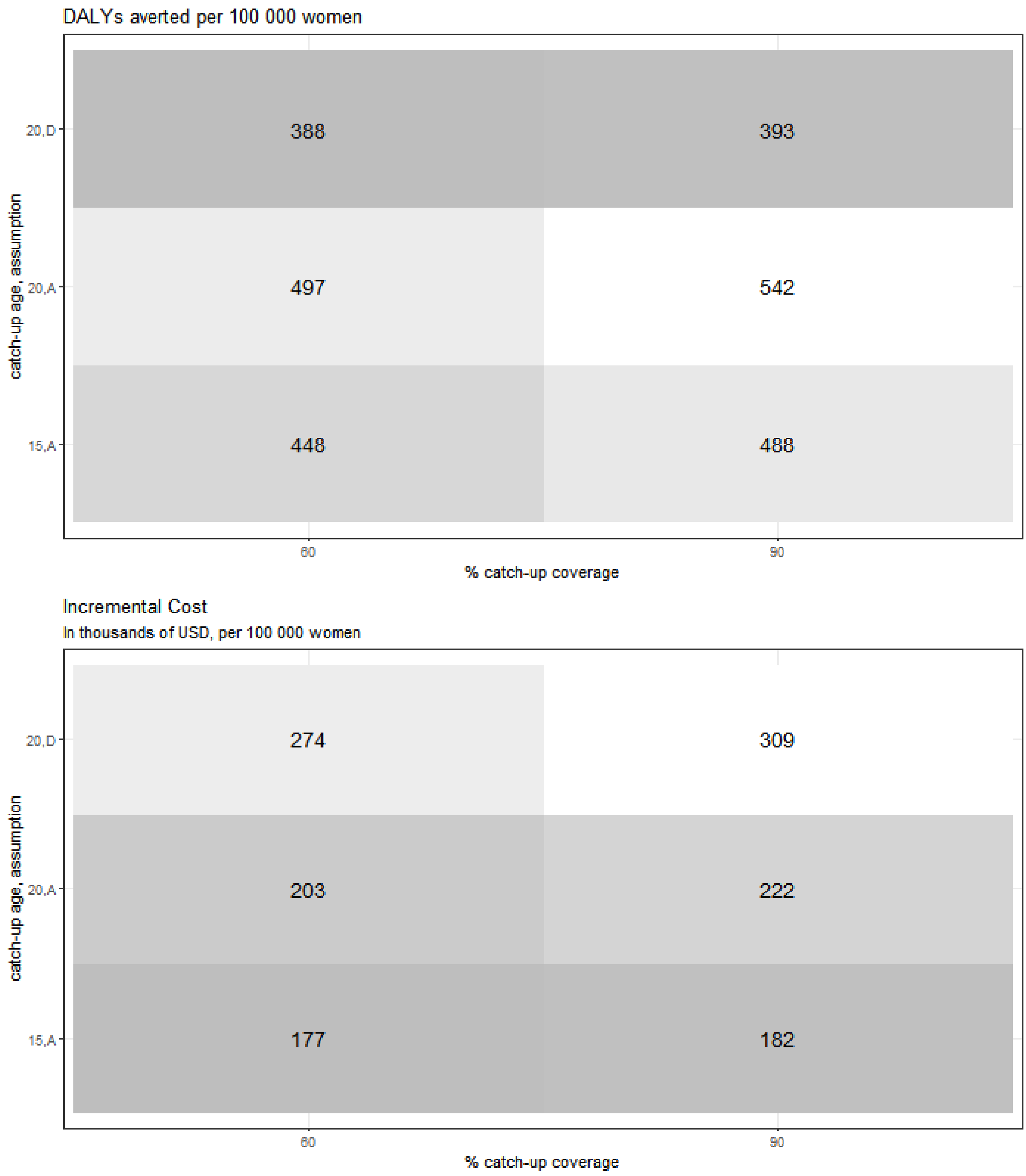

[a] Costs are given in $USD at 2020 prices. Costs and disability-adjusted life years (DALYs) averted are discounted at 3%. See Figure 3 for the corresponding ICER.

#### Section 3: Results in $IUSD

We present our results in USD, since the main goal of this analysis is to inform Indian health officials' decisions about cervical cancer prevention in India. This results in higher ICERs than in studies that present results in IUSD, due to the purchasing power parity conversion rate from INR to IUSD used for non-tradable goods, which makes cervical cancer treatment costs (e.g. salary of medical staff) relatively more expensive than tradable goods (e.g. vaccine dose).

**Figure B.10: Undiscounted incremental costs and health effects of single- and two-dose vaccination versus no vaccination ($IUSD).[a]**

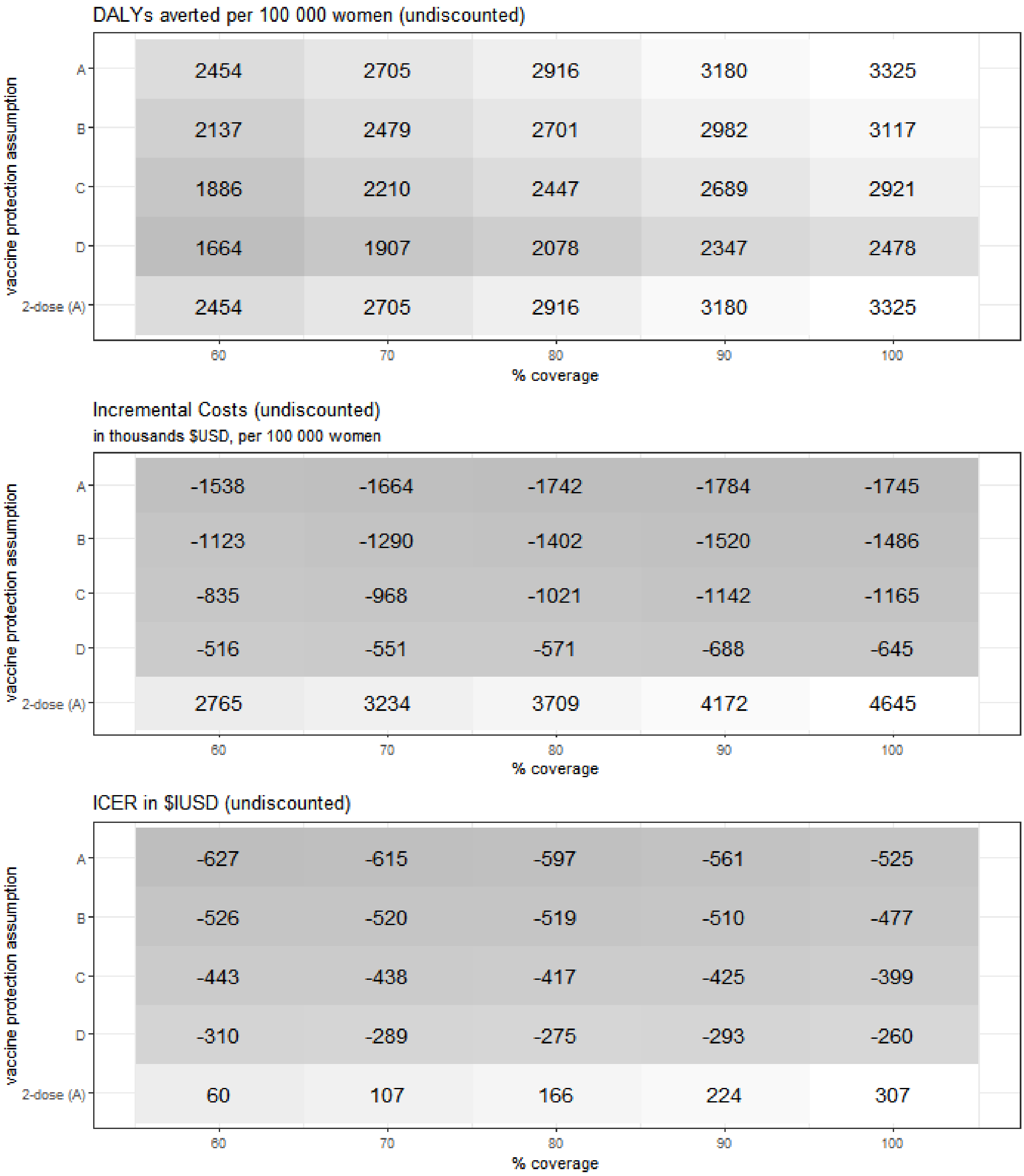

[a] Costs are given in $IUSD at 2020 prices. Costs and disability-adjusted life years (DALYs) averted are not discounted. ICER denotes incremental cost-effectiveness ratio per DALY averted relative to no vaccination.

**Figure B.11: Incremental costs and health effects of single- and two-dose vaccination versus no vaccination.**

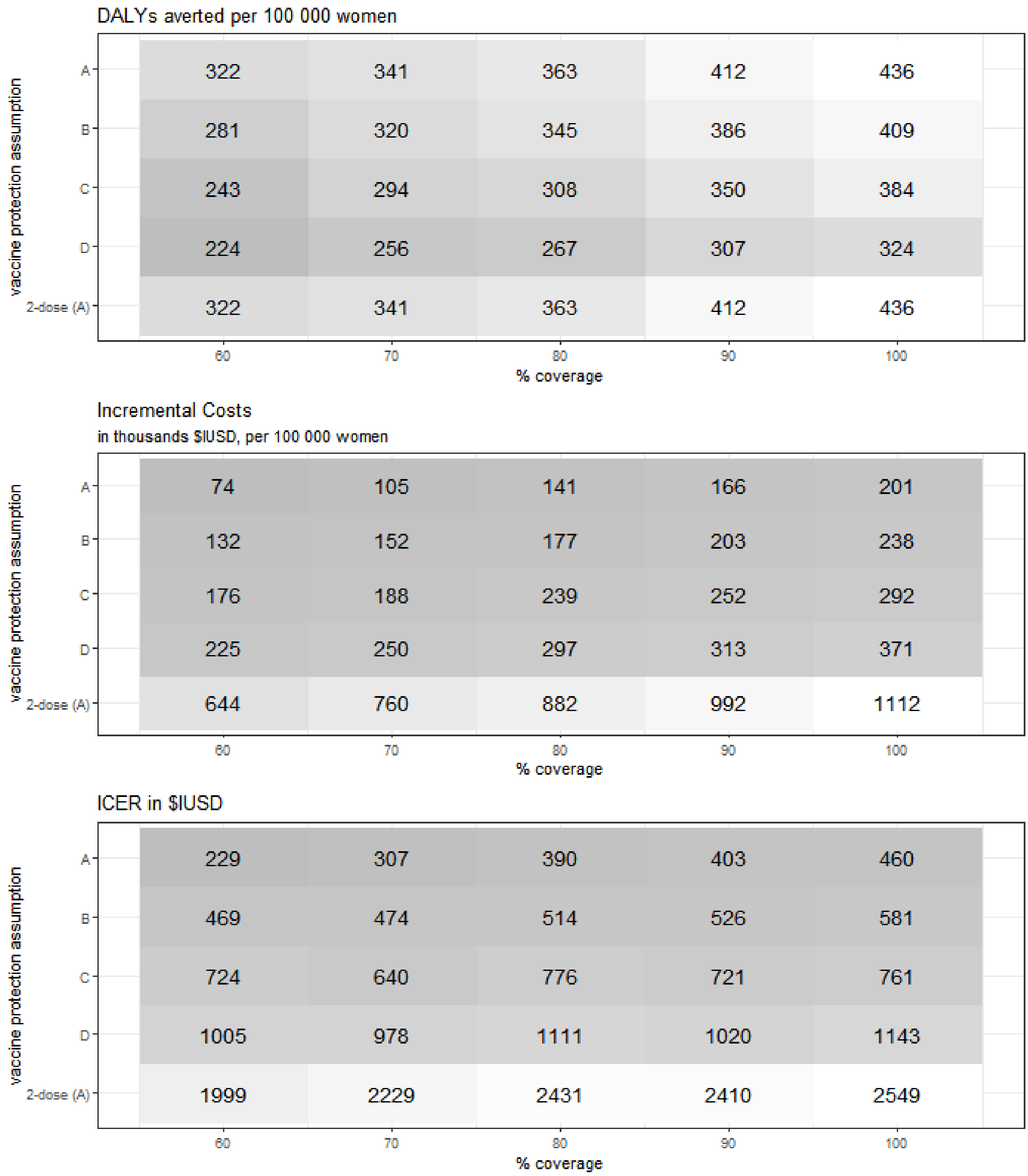

[a] Costs are given in $IUSD at 2020 prices. Costs and disability-adjusted life years (DALYs) averted are discounted at 3%. ICER denotes incremental cost-effectiveness ratio per DALY averted relative to no vaccination. We consider two cost-effectiveness thresholds for the ICER, a) 100% of Indian GDP per capita, as per WHO recommendation ($IUSD 6504), and b) 30% of Indian GDP per capita ($IUSD 1951).

**Figure B.12: Incremental costs and health effects of two-dose versus single-dose vaccination (assuming lifetime protection for two-dose vaccination).[a]**

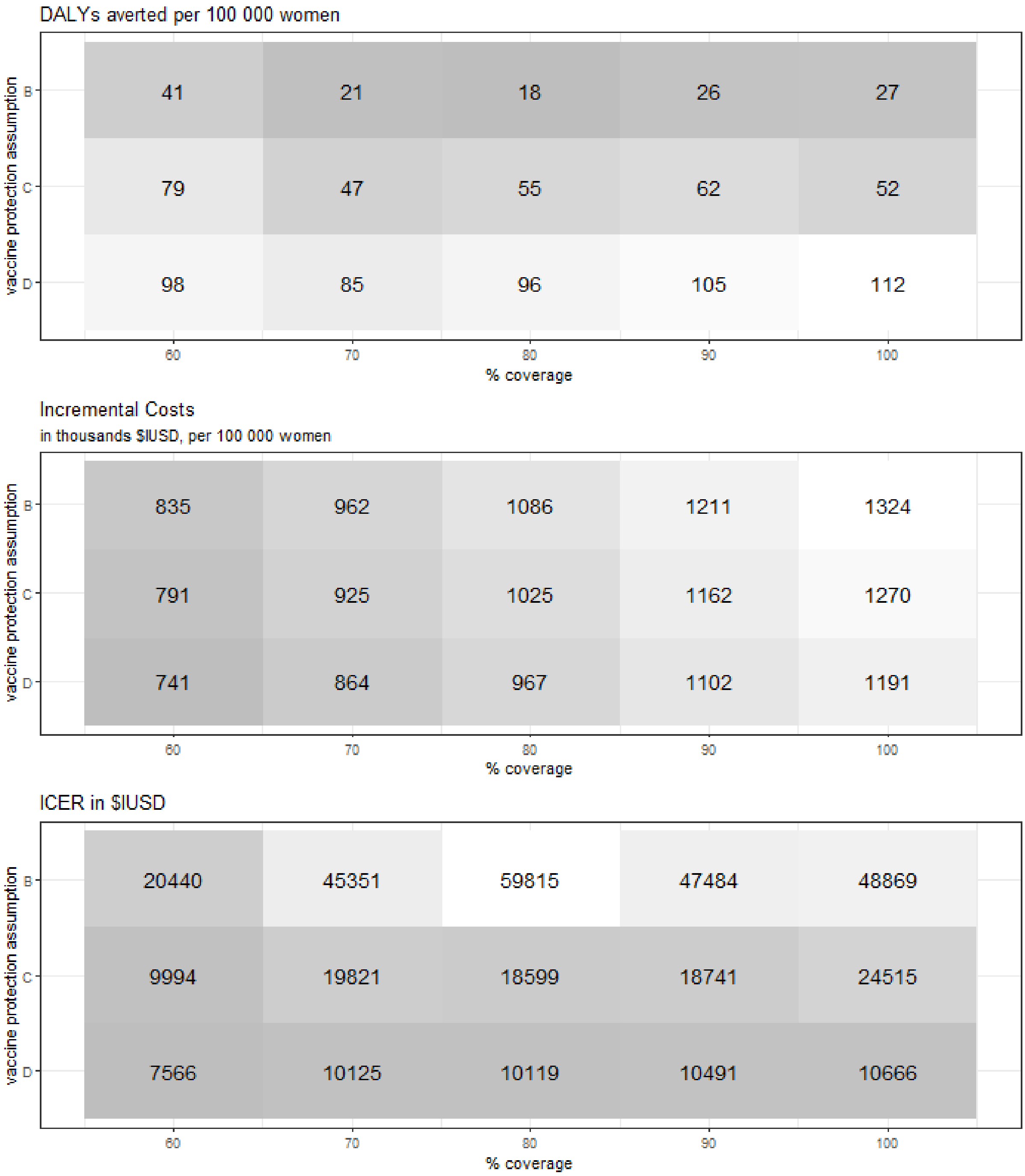

[a] Costs are given in $IUSD at 2020 prices. Costs and disability-adjusted life years (DALYs) averted are discounted at 3%. ICER denotes incremental cost-effectiveness ratio per DALY averted relative to no vaccination. We consider two cost-effectiveness thresholds for the ICER, a) 100% of Indian GDP per capita, as per WHO recommendation ($IUSD 6504), and b) 30% of Indian GDP per capita ($IUSD 1951).

**Figure B.13: Health economic outcomes by catch-up scenario.**[a]

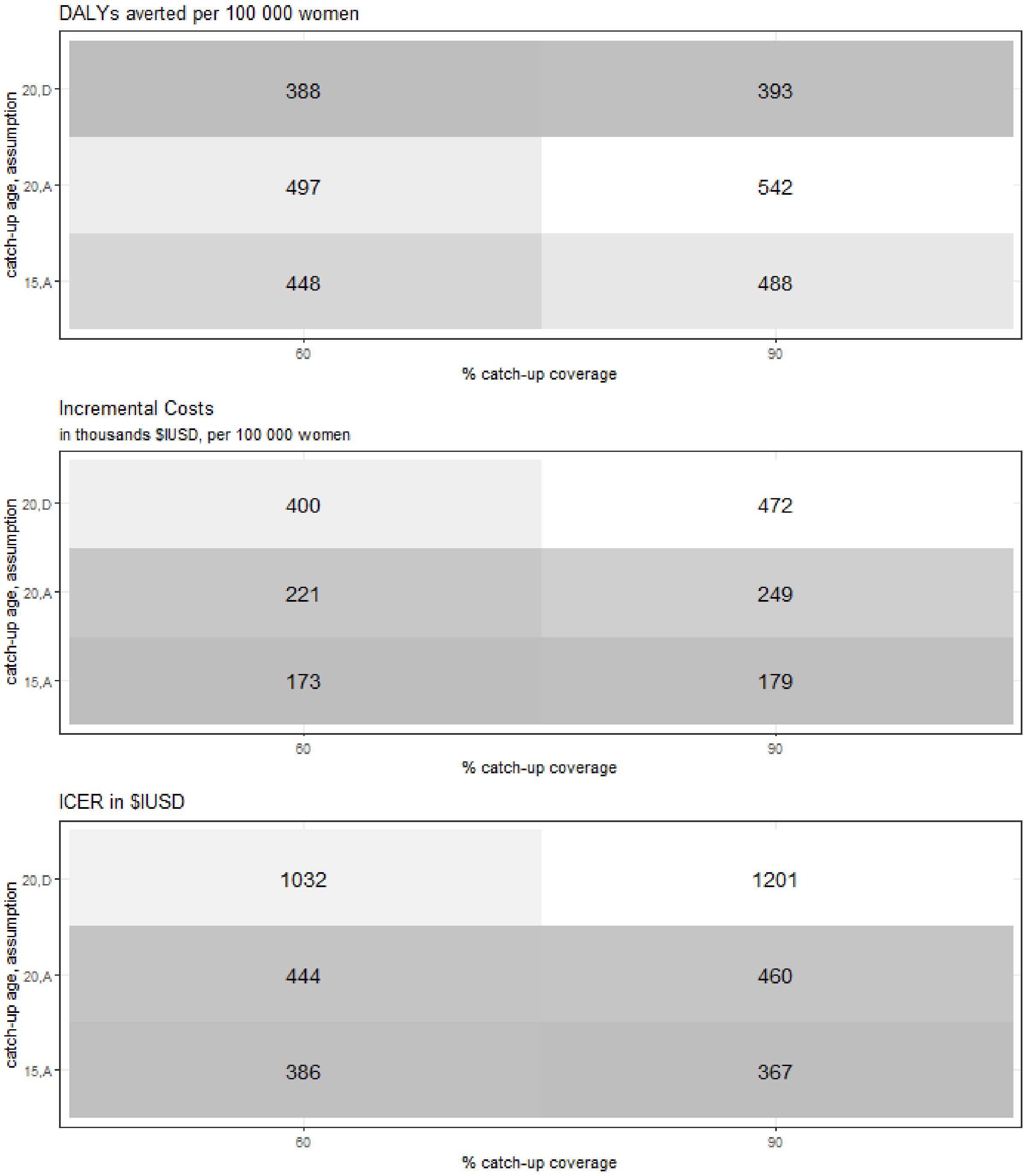

[a] Costs are given in $IUSD at 2020 prices. Costs and disability-adjusted life years (DALYs) averted are discounted at 3%. ICER denotes incremental cost-effectiveness ratio per DALY averted relative to no vaccination. We consider two cost-effectiveness thresholds for the ICER, a) 100% of Indian GDP per capita, as per WHO recommendation ($IUSD 6504), and b) 30% of Indian GDP per capita ($IUSD 1951).

**Figure B.14: Univariate sensitivity analysis on the ICER for cost variables.**

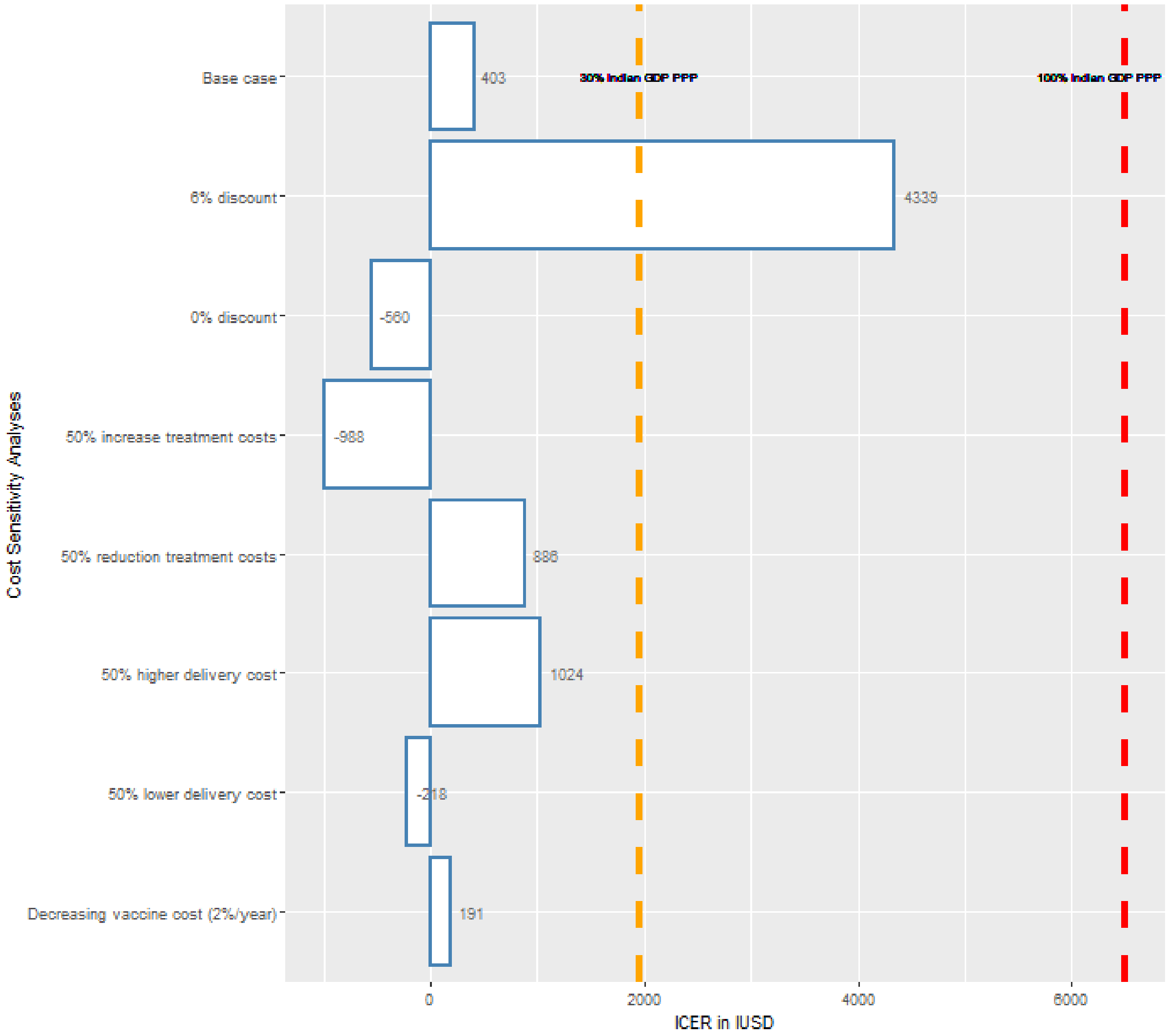

[a] Costs are given $IUSD at 2020 prices. Costs and disability-adjusted life years (DALYs) averted are discounted at 3%. ICER denotes incremental cost-effectiveness ratio. The dashed lines denote the range for cost-effectiveness thresholds, 30% of Indian GDP per capita in $IUSD (orange) and 100% of Indian GDP per capita in $IUSD (red). PPP=purchasing power parity.
